# Diffusion-weighted magnetic resonance imaging for the diagnosis of giant cell arteritis – a comparison with T1-weighted black-blood imaging

**DOI:** 10.1101/2023.07.14.23288940

**Authors:** Luca Seitz, Susana Bucher, Lukas Bütikofer, Britta Maurer, Harald M. Bonel, Franca Wagner, Fabian Lötscher, Pascal Seitz

## Abstract

**Objectives:** To investigate the diagnostic performance of diffusion-weighted imaging (DWI) of the superficial cranial arteries in the diagnosis of giant cell arteritis (GCA).

**Methods:** Retrospectively, 156 patients with clinically suspected GCA were included. A new 4-point ordinal DWI rating scale was developed. A post-contrast, fat-suppressed, T1-weighted “black-blood” sequence (T1-BB) was rated for comparison. Ten arterial segments were assessed: common superficial temporal arteries, temporal and parietal branches, occipital and posterior auricular arteries bilaterally. The expert clinical diagnosis after ≥ 6 months of follow-up was the diagnostic reference standard. Diagnostic accuracy was evaluated for different rating methods.

**Results:** The study cohort consisted of 87 patients with and 69 without GCA. For DWI, the area under the curve was 0.90. For a cut-off of ≥ 2 consecutive pathological slices, DWI showed a sensitivity of 75.9%, a specificity of 94.2% and a positive likelihood ratio of 13.09. With a cut-off of ≥ 3 consecutive pathological slices, sensitivity was 70.1%, specificity was 98.6%, and the positive likelihood ratio was 48.38. For the T1-BB, values were 88.5%, 88.4% and 7.63, respectively. The inter-rater analysis for DWI with a cut-off of ≥ 2 pathological slices showed a kappa of 1.00 on the patient level and 0.85 on the arterial segment level. For the T1-BB the kappa was 0.78 and 0.79, respectively.

**Conclusion:** DWI of the superficial cranial arteries demonstrates a good diagnostic accuracy and reliability for the diagnosis of GCA. DWI is widely available and can be used immediately in clinical practice for patients with suspected GCA.

## Introduction

Giant cell arteritis (GCA) typically affects the superficial cranial arteries (SCAs) [1]. Early diagnosis and initiation of treatment are critical [2,3]. Confirmation of diagnosis by imaging or biopsy is advised [4,5]. The use of a 3-Tesla, contrast-enhanced, high-resolution, fat-suppressed T1-weighted, spin-echo sequence (T1-black-blood (T1-BB)) is recommended for magnetic resonance imaging (MRI) of the SCAs [6]. The T1-BB is the current reference sequence for the diagnosis of GCA [6,7,8]. Potential disadvantages include its long acquisition time and limited availability. Also, the T1-BB sequence is performed only when vasculitis is suspected. Especially for headache, this is often not the case and a standard MRI protocol without sequences for vasculitis is performed instead [6,8]. Such protocols are currently considered insufficient to detect GCA.

GCA would ideally be detectable with a standard head MRI protocol. In 2018, Ironi et al. published on diffusion-weighted imaging (DWI) in aortitis [9]. In 2019, Matsuoka et al. reported, to our knowledge, the first case with an abnormal DWI signal of the temporal arteries (TAs) [10]. Recently, a few additional cases of DWI in GCA were published [11–13]. DWI is based on the measurement of random Brownian motion of water molecules. Highly cellular tissues and tissues with cellular oedema have restricted diffusion and appear as relatively hyperintense regions in DWI, more pronounced at high b-values [14–18]. The DWI sequence is acquired quickly, does not require intravenous contrast material, and is included in most head MRI protocols.

This study investigates how SCAs can be evaluated with DWI in suspected GCA and how the diagnostic performance of DWI compares to T1-BB, the established reference MRI sequence for this indication. To address these questions, we conducted a study of 156 patients with clinically suspected GCA.

## Methods

This is a retrospective, monocentric study at the University Hospital Bern, Switzerland, a tertiary vasculitis referral centre. The study is in accordance with the Declaration of Helsinki and was approved by the ethics committee Bern, Switzerland, in December 2021 (ID: 2021-02169). This manuscript is in accordance with the “Standards for Reporting of Diagnostic Accuracy Studies” (STARD) [19].

### Study population

Inclusion criteria: Age ≥ 50 years; evaluation for clinically suspected GCA (any reason) or suspected relapse of GCA; available head MRI scan at the time of evaluation (performed between January 1^st^, 2018, and December 31^st^, 2021); written informed consent. Exclusion criteria (≥ one had to be met): severe image artifacts; DWI or T1-BB sequences not performed; vasculitis other than GCA; complete clinical and laboratory remission for less than four months after treatment discontinuation (only for relapses). Hospital records were searched, and 208 consecutive patients were identified. Fifty-two patients were excluded (21 GCA, 31 non-GCA): 2 with a missing DWI sequence, 35 with a missing T1-BB sequence, 1 with MR artifacts, 4 with non-GCA vasculitis, and 10 relapses (**Supplementary Figure S1**). The pre-specified diagnostic reference standard was the clinical expert diagnosis more than six months after the initial diagnosis. It was established by two vasculitis experts (LS, PS, or FL; senior rheumatologists) through comprehensive analysis of the medical records prior to image evaluation. For 2/156 patients, the diagnosis differed between experts (polymyalgia rheumatica vs. polyarthritis) and was determined as polyarthritis by consensus.

### Image acquisition

Imaging was performed on 3-Tesla scanners (Prisma Fit, Skyra Fit, Verio or Vida; Siemens, Erlangen, Germany) with a dedicated head and neck coil with 20 or 64 channels. Slices were aligned along the skull base, acquired in the axial plane, and covered the distance from the hard palate to the vertex for all sequences. The 3D arterial time-of-flight MR angiography (3D-TOF-MRA) had a slice thickness of 0.5mm. A pre-contrast, fat-saturated T2-weighted spin-echo sequence (T2-fs), was acquired with: slice thickness 3 - 4 mm; acquisition matrix 348 - 406 × 384 – 448; field of view (FOV) 199 × 220 mm; TR median 4790 ms (interquartile range (IQR) 4435 – 4790 ms); TE median 106 ms (IQR 103 – 116 ms). The DWI sequence was acquired with: b-value of 0 and 1000 s/mm^2^ in 100%; slice thickness 4mm in 98.7%; acquisition matrix 192 × 192 in 93.6%; FOV 220 × 220 mm in 98.7%; voxel size 1.15 × 1.15 × 4.00 mm in 93.6%; TR median 5750 ms (IQR 4570 - 5750 ms); TE median 61 ms (IQR 61 - 67 ms); flip angle 180 in 98.7%; readout-segmented multi-shot echo-planar imaging (EPI) (Resolve) in 98.7% of cases [20]. **Supplementary Table S1** shows more detailed DWI parameters. The spatial resolution of the corresponding apparent diffusion-coefficient (ADC) images was too low to reliably detect SCAs and thus was not analysed. The T1-BB was acquired after intravenous injection of a gadolinium-based contrast agent in three consecutive blocks of ten slices with: slice thickness 3 mm; slice spacing 6 mm; acquisition matrix 1024 × 768; FOV 200 × 200 mm; voxel size 0.260 × 0.195 × 3 mm; TR 500 ms; TE 22 ms [21].

### Image evaluation

Images were read with Sectra IDS7 (DICOM software, version 23.1) by LS (146 patients) and PS (10 patients and 20 MRI scans for inter-reader analysis), senior rheumatologists with specialization in vasculitis imaging and 12 and 11 years of work experience, respectively. Readers were blinded to the reference diagnosis and all clinical information apart from sex and age. The 20 scans for inter-reader analysis were randomly chosen from patients who had an SCA biopsy. The whole depicted length of ten arterial segments was assessed: common superficial TA (CSTA), frontal and parietal branches of the TA, posterior auricular artery, occipital artery. First, the DWI images were rated. If necessary, the brightness was adjusted. **Supplementary Figure S2** shows an ideal brightness setting. The CSTAs were only rated once they entered the subcutaneous tissue, i.e., the segment that runs parallel to the ear canal was not rated because it is not possible to differentiate it from surrounding structures with DWI. In a next step, the T2-fs images were rated at the same location of the DWI rating (corresponding slice, using crosshair). Only then, the T1-BB was rated. Once rating of the T2-fs was started, DWI ratings were not changed.

### Rating of arteries

The 3D-TOF-MRA was used to identify arteries. The T1-BB sequence was rated according to a published scale: 0 = no mural thickening and no mural enhancement; 1 = no mural thickening with slight mural enhancement; 2 = mural thickening with prominent mural enhancement; 3 = strong mural thickening with strong mural and perivascular enhancement [21–24]. T1-BB scores 0 and 1 are considered physiological and scores 2 and 3 are considered to represent vessel wall inflammation [21–24]. New rating scales were defined for the T2-fs and DWI sequences after they were tested in daily practice by LS, PS and FL. In T2-fs images, arteries were rated according to the following scale: 0 = vessel wall not visible; 1 = vessel wall visible; 2 = vessel wall prominently visible; 3 = vessel wall prominently visible and perivascular oedema. T2-fs scores 0 and 1 are considered physiological, score 2 as borderline (either physiological or vascular inflammation) and score 3 is considered to represent vessel wall inflammation. **Supplementary Figure S3** shows the T2-fs-scoring. For DWI (b-value of 1000 s/mm^2^), a simplified scale was defined because vessel wall and lumen are usually indistinguishable: 0 = artery not visible; 1 = artery slightly visible; 2 = artery prominently visible; 3 = artery brightly visible (**Figure 1**). The spatial resolution of DWI is low (> 1mm) and the anatomical correlation between sequences when using the crosshair can vary by a few millimetres. Accordingly, there is a risk of misidentification of non-arterial structures, especially for the CSTAs and occipital arteries, where adjacent lymph nodes, parts of the parotid gland or veins may look similar [14,25]. Therefore, three different DWI scales were evaluated. For DWI version 1 (DWI-1), maximum DWI score present over ≥ 1 slice, and DWI version 2 (DWI-2), maximum DWI score present over ≥ 2 slices, the following interpretation was pre-specified: DWI scores 0 and 1 are considered physiological, scores 2 and 3 are considered to represent vessel wall inflammation. For DWI version 3 (DWI-3), the number of consecutive slices with a DWI score of 2 *or* 3 was counted for every arterial segment (segments with ≥ 7 slices were combined). For DWI-3, no cut-off was pre-specified. The 20 MRI scans for the inter-rater analysis were assessed for DWI-2 and T1-BB. Examples of DWI scores, pitfalls of DWI and T1-BB-imaging, and cases with follow-up imaging are provided in a **Supplementary Atlas**.

**Figure 1.**
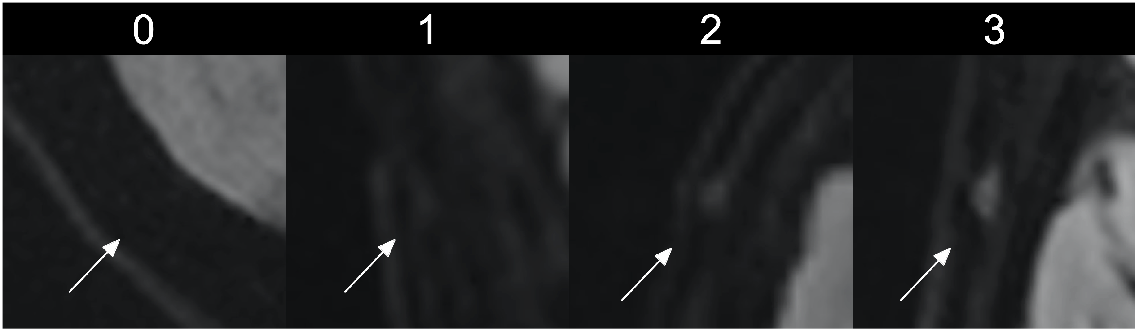
DWI-scoring for the superficial cranial arteries. DWI-scoring (0 – 3): 0 = artery not visible; 1 = artery slightly visible; 2 = artery prominently visible; 3 = artery brightly visible. (Figure 1 without arrows is provided as supplementary Figure S4)

### Statistics

Statistical analyses were done with Stata; figures were made with R [26,27]. Patient characteristics are reported as absolute and relative frequencies for categorical variables and as median with IQR for continuous variables. They were compared between groups using Fisher’s exact and the Mann-Whitney-Wilcoxon test for categorical and continuous variables, respectively. The proportion of correct classifications, sensitivity, and specificity are reported as absolute and relative frequencies with Wilson 95%-confidence interval (CI). Sensitivity and specificity were compared between methods using the McNemar’s test. Binary agreement at the patient or segment level were quantified by Cohen’s kappa with an analytical 95%-CI. Segment-level correlation was quantified by Spearman’s Rho with 95%-CI (based on Fisher transformation). The area under the curve (AUC) of the receiver-operating-characteristic (ROC) is reported with asymptotic DeLong 95%-CI. Best cut-off was determined using the methods of Liu and Youden [28,29].

## Results

Data from 156 patients were analysed, 87 (55.8%) with GCA and 69 (44.2%) with different diagnoses, listed in **Supplementary Table S2**. A total of 151 (96.8%) patients were evaluated for suspected new-onset GCA and 5 (3.2%) for suspected relapsing GCA. Cranial manifestations were present in 128 (82.1%) patients and 28 (17.9%) had only non-cranial manifestations. **Table 1** shows the patients’ characteristics [30,31].

**Table 1.** Patients’ characteristics. ^a^, n (%) unless stated otherwise; ^b^, median (IQR); ^c^, established atherosclerotic comorbidity (e.g., myocardial infarction); ^d^, persistent vision loss (complete or incomplete, unilateral or bilateral); ^e^, ≥ 1 dose of methylprednisolone (500 – 1000 mg) was given intravenously ≥ 1 day prior to the MRI; ^f^, classification criteria are not met by definition if vasculitis is not present; ^g^, was not assessed for non-GCA cases. Missing values (Diplopia – 1; Abnormal TA exam – 8; Thickening of TA – 14; ESR – 15; Anaemia/Thrombocytosis – 1; Duration of CS use before MRI – 1). n.a.: not applicable; TA: temporal artery.

| Characteristic <sup>a</sup> | Total study population |  |  |  | Patients with cranial manifestations |  |  |  |
| --- | --- | --- | --- | --- | --- | --- | --- | --- |
|  | Total<br>(N = 156) | No GCA<br>(N = 69) | GCA<br>(N = 87) | P-Value | Total<br>(N = 128) | No GCA<br>(N = 53) | GCA<br>(N = 75) | P-value |
| Age (years) <sup>b</sup> | 71 (65 - 77) | 69 (62 - 76) | 72 (67 - 77) | 0.038 | 72 (66 - 77) | 69 (63 - 76) | 73 (68 - 78) | 0.032 |
| Female patients | 92 (59.0%) | 36 (52.2%) | 56 (64.4%) | 0.14 | 81 (63.3%) | 31 (58.5%) | 50 (66.7%) | 0.36 |
| 1990 - ACR criteria | 70 (44.9%) | n.a. <sup>f</sup> | 70 (80.5%) | n.a. | 66 (51.6%) | n.a. <sup>f</sup> | 66 (88.0%) | n.a. |
| 2022 - ACR/EULAR criteria | 84 (53.8%) | n.a. <sup>f</sup> | 84 (96.6%) | n.a. | 74 (57.8%) | n.a. <sup>f</sup> | 74 (98.7%) | n.a. |
| 2022 - ACR/EULAR criteria (points) <sup>b</sup> | 10 (6 - 13) | 6 (4 - 9) | 13 (10 - 15) | <0.001 | 11 (7 - 14) | 7 (4 - 9) | 13 (11 - 15) | <0.001 |
| Atherosclerosis <sup>c</sup> | 42 (26.9%) | 21 (30.4%) | 21 (24.1%) | 0.47 | 37 (28.9%) | 18 (34.0%) | 19 (25.3%) | 0.33 |
| Constitutional symptoms | 98 (62.8%) | 37 (53.6%) | 61 (70.1%) | 0.045 | 76 (59.4%) | 24 (45.3%) | 52 (69.3%) | 0.01 |
| New-onset headache | 102 (65.4%) | 38 (55.1%) | 64 (73.4%) | 0.018 | 102 (79.7%) | 38 (71.7%) | 64 (85.3%) | 0.08 |
| Scalp tenderness | 47 (30.1%) | 19 (27.5%) | 28 (32.2%) | 0.60 | 47 (36.7%) | 19 (35.8%) | 28 (37.3%) | 1.0 |
| Jaw claudication | 39 (25.0%) | 7 (10.1%) | 32 (36.8%) | <0.001 | 39 (30.5%) | 7 (13.2%) | 32 (42.7%) | <0.001 |
| Diplopia | 17 (10.9%) | 6 (8.6%) | 11 (12.6%) | 0.15 | 17 (13.3%) | 6 (11.4%) | 11 (14.6%) | 0.18 |
| Vision loss <sup>d</sup> | 28 (17.9%) | 13 (18.8%) | 15 (17.2%) | 0.21 | 28 (21.9%) | 13 (24.5%) | 15 (20.0%) | 0.22 |
| PMR symptoms | 85 (54.5%) | 42 (60.9%) | 43 (49.4%) | 0.20 | 64 (50.0%) | 28 (52.8%) | 36 (48.0%) | 0.72 |
| Abnormal TA exam | 61 (39.1%) | 17 (24.6%) | 44 (50.6%) | 0.003 | 61 (47.7%) | 17 (32.1%) | 44 (58.7%) | 0.008 |
| Thickening of TA | 26 (16.7%) | 4 (5.8%) | 22 (25.3%) | 0.010 | 26 (20.3%) | 4 (7.5%) | 22 (29.3%) | 0.021 |
| CRP (mg/L) <sup>b</sup> | 63 (24 - 123) | 53 (5 - 111) | 78 (42 - 130) | 0.017 | 60 (15 - 109) | 17 (4 - 68) | 78 (39 - 130) | <0.001 |
| ESR (mm/1h) <sup>b</sup> | 71 (38 - 86) | 51 (28 - 77) | 80 (50 - 89) | <0.001 | 67 (30 - 84) | 42 (24 - 69) | 78 (44 - 86) | <0.001 |
| Anaemia | 86 (55.1%) | 29 (42.0%) | 57 (65.5%) | 0.004 | 66 (51.6%) | 19 (35.8%) | 47 (62.7%) | 0.004 |
| Thrombocytosis | 24 (15.4%) | 8 (11.6%) | 16 (18.4%) | 0.27 | 17 (13.3%) | 3 (5.7%) | 14 (18.7%) | 0.037 |
| CS started before MRI | 54 (34.6%) | 25 (36.2%) | 29 (33.3%) | 0.74 | 47 (36.7%) | 21 (39.6%) | 26 (34.7%) | 0.58 |
| Duration of CS use before MRI (days) <sup>b</sup> | 0 (0 - 2) | 0 (0 - 24) | 0 (0 - 1) | 0.014 | 0 (0 - 2) | 0 (0 - 24) | 0 (0 - 1) | 0.031 |
| CS i.v. before MRI <sup>e</sup> | 21 (13.5%) | 5 (7.2%) | 16 (18.4%) | 0.06 | 19 (14.8%) | 5 (9.4%) | 14 (18.7%) | 0.21 |
| TA biopsy performed | 93 (59.6%) | 24 (34.8%) | 69 (79.3%) | <0.001 | 79 (61.7%) | 17 (32.1%) | 62 (82.7%) | <0.001 |
| TA histology vasculitis | 52 (33.3%) | 0 (0%) | 52 (59.8%) | <0.001 | 47 (36.7%) | 0 (0%) | 47 (62.7%) | <0.001 |
| Imaging with LVV | n.a. <sup>g</sup> | n.a. <sup>g</sup> | 58 (66.7%) | n.a. | n.a. <sup>g</sup> | n.a. <sup>g</sup> | 49 (65.3%) | n.a. |

Scoring methods DWI-1, DWI-2, and T1-BB were each analysed for the subgroup with cranial manifestations and the total study population. Methods DWI-3 and T2-fs were solely analysed for the latter. **Table 2** shows the measures of diagnostic accuracy of DWI-1, DWI-2, and T1-BB for both groups compared to the reference diagnosis. While sensitivities for DWI-2 and T1-BB were higher for patients with cranial manifestations compared with the total study population, specificity was almost identical for DWI-2 and only slightly lower for T1-BB. More detailed information regarding the proportion of correct diagnoses for DWI-1, DWI-2 and T1-BB compared to the reference diagnosis are provided in **Supplementary Table S3. Supplementary Table S4** displays the comparison of the binary classification (normal vs. pathological) of DWI-2 with T1-BB at the segment level, and **Supplementary Table S5** shows the sensitivity and specificity of DWI-2 with T1-BB as reference for the segment level.

**Table 2.** Measures of diagnostic accuracy for DWI-1, DWI-2 and T1-BB methods compared to the reference diagnosis. ^a^, % (95%-confidence interval); ^b^, n (%, 95%-confidence interval). LR: likelihood ratio; DWI-1: diffusion-weighted imaging with maximum score over ≥ 1 slice; DWI-2: diffusion-weighted imaging with maximum score present over ≥ 2 consecutive slices; T1-BB: T1-black-blood.

|  | Abnormal test / GCA | Sensitivity <sup>a</sup> | P-value (vs. T1-BB) | Normal test / No GCA | Specificity <sup>a</sup> | P-value (vs. T1-BB) | Positive LR <sup>a</sup> | Negative LR <sup>a</sup> | Correct diagnosis <sup>b</sup> |
| --- | --- | --- | --- | --- | --- | --- | --- | --- | --- |
| **Total study population (n = 156)** |  |  |  |  |  |  |  |  |  |
| **DWI-1** | 85/87 | 97.7%<br>(92.0 – 99.4%) | 0.011 | 26/69 | 37.7%<br>(27.2 – 49.5%) | <0.001 | 1.57<br>(1.30 – 1.89) | 0.06<br>(0.01 – 0.25) | 111 (71.2%,<br>63.6 – 77.7%) |
| **DWI-2** | 66/87 | 75.9%<br>(65.9 – 83.6%) | <0.001 | 65/69 | 94.2%<br>(86.0 – 97.7%) | 0.10 | 13.09<br>(5.02 – 34.13) | 0.26<br>(0.18 – 0.37) | 131 (84.0%,<br>77.4 – 88.9%) |
| **T1-BB** | 77/87 | 88.5%<br>(80.1 – 93.6%) |  | 61/69 | 88.4%<br>(78.8 – 94.0%) |  | 7.63<br>(3.96 – 14.71) | 0.13<br>(0.07 – 0.23) | 138 (88.5%,<br>82.5 – 92.6%) |
| **Patients with cranial manifestations (n = 128)** |  |  |  |  |  |  |  |  |  |
| **DWI-1** | 73/75 | 97.3%<br>(90.8 – 99.3%) | 0.18 | 19/53 | 35.8%<br>(24.3 – 49.3%) | <0.001 | 1.52<br>(1.24 – 1.86) | 0.07<br>(0.02 – 0.31) | 92 (71.9%,<br>63.5 – 78.9%) |
| **DWI-2** | 61/75 | 81.3%<br>(71.1 – 88.5%) | 0.003 | 50/53 | 94.3%<br>(84.6 – 98.1%) | 0.16 | 14.37<br>(4.76 – 43.36) | 0.20<br>(0.12 – 0.32) | 111 (86.7%,<br>79.8 – 91.5%) |
| **T1-BB** | 70/75 | 93.3%<br>(85.3 – 97.1%) |  | 48/53 | 90.6%<br>(79.7 – 95.9%) |  | 9.89<br>(4.29 – 22.83) | 0.07<br>(0.03 – 0.17) | 118 (92.2%,<br>86.2 – 95.7%) |

DWI-3 was evaluated to determine the number of consecutive slices for a single arterial segment at which a diagnosis of GCA can be made with a very high degree of certainty. Liu’s method and Youden’s index both reported ≥ 2 as the optimal cut-off for the number of slices with a DWI score of 2 *or* 3. **Figure 2** shows the ROC curve for DWI-3. **Table 3** shows the sensitivities and specificities at different cut-points for DWI-3 based on the maximum number of pathological slices per patient. If there is ≥ 1 segment with ≥ 3 consecutive slices with a DWI score of 2 *or* 3, the specificity for the diagnosis of GCA was 98.6% (95%-CI 92.2 - 99.7%) with a positive likelihood ratio of 48.38 (95%-CI 6.88 - 340.23).

**Table 3.** Measures of diagnostic accuracy for each cut-point of DWI-3 compared to the reference diagnosis. Based on the total study population. Optimal cut-point: Liu ≥ 2 slices; Youden ≥ 2 slices. ^a^, % (95%-confidence interval); ^b^, includes cases with more than 7 slices. n.a.: not applicable; LR: likelihood ratio.

| Cut-point | Sensitivity <sup>a</sup> | Specificity <sup>a</sup> | Correctly classified <sup>a</sup> | Positive LR <sup>a</sup> | Negative LR <sup>a</sup> |
| --- | --- | --- | --- | --- | --- |
| ≥ 0 | 100.0% (95.8 – 100.0%) | 0.0% (0.0 – 5.3%) | 55.8% (47.9 – 63.3%) | 1.00 | n.a. |
| ≥ 1 | 97.7% (92.0 – 99.4%) | 37.7% (27.2 – 49.5%) | 71.2% (63.6 – 77.7%) | 1.57 (1.30 – 1.89) | 0.06 (0.01 – 0.25) |
| ≥ 2 | 75.9% (65.9 – 83.6%) | 94.2% (86.0 – 97.7%) | 84.0% (77.4 – 88.9%) | 13.09 (5.02 – 34.13) | 0.26 (0.18 – 0.37) |
| ≥ 3 | 70.1% (59.8 – 78.7%) | 98.6% (92.2 – 99.7%) | 82.7% (76.0 – 87.8%) | 48.38 (6.88 – 340.23) | 0.30 (0.22 – 0.42) |
| ≥ 4 | 65.5% (55.1 – 74.7%) | 98.6% (92.2 – 99.7%) | 80.1% (73.2 – 85.6%) | 45.21 (6.42 – 318.28) | 0.35 (0.26 – 0.47) |
| ≥ 5 | 54.0% (43.6 – 64.1%) | 100.0% (94.7 – 100.0%) | 74.4% (67.0 – 80.6%) | n.a. | 0.46 (0.37 – 0.58) |
| ≥ 6 | 47.1% (37.0 – 57.5%) | 100.0% (94.7 – 100.0%) | 70.5% (62.9 – 77.1%) | n.a. | 0.53 (0.43 – 0.64) |
| ≥ 7<sup>b</sup> | 42.5% (32.7 – 53.0%) | 100.0% (94.7 – 100.0%) | 67.9% (60.3 – 74.8%) | n.a. | 0.57 (0.48 – 0.69) |
| > 7<sup>b</sup> | 0.0% (0.0 – 4.2%) | 100.0% (94.7 – 100.0%) | 44.2% (36.7 – 52.1%) | n.a. | 1.00 |

**Figure 2.**
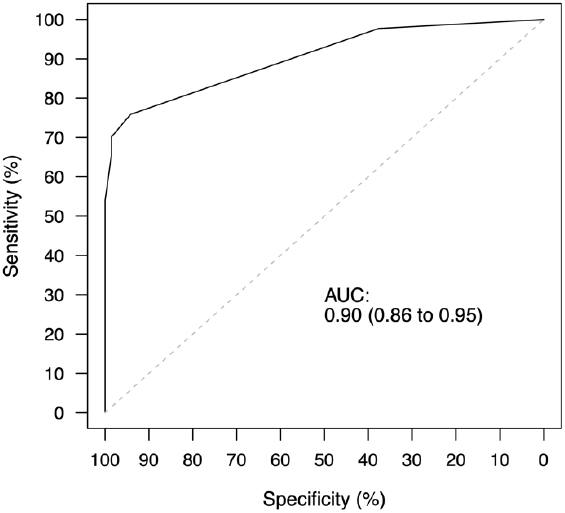
ROC curve for the maximum number of consecutive pathological slices per patient (DWI-3). Compared to the reference diagnosis and based on 1327 arterial segments from 156 patients (total study population). Area under the curve (AUC): 0.90 (95%-confidence interval 0.86 – 0.95)

The difficult assessment of the CSTAs is supported by the lower kappa for the CSTAs in the inter-rater analysis compared to the overall kappa: Left CSTA (0.59, 95%-CI 0.08 - 1.0), right CSTA (0.64, 95%-CI 0.27 – 1.00), overall (0.85, 95%-CI 0.76 – 0.93). In addition, occipital lymph nodes were found in 93% and the posterior auricular arteries were frequently not detectable at all on DWI images. DWI-2x was thus evaluated as an exploratory analysis, considering only the DWI-2 results from the more reliably assessable parietal and frontal branches (**Table 4**).

**Table 4.**
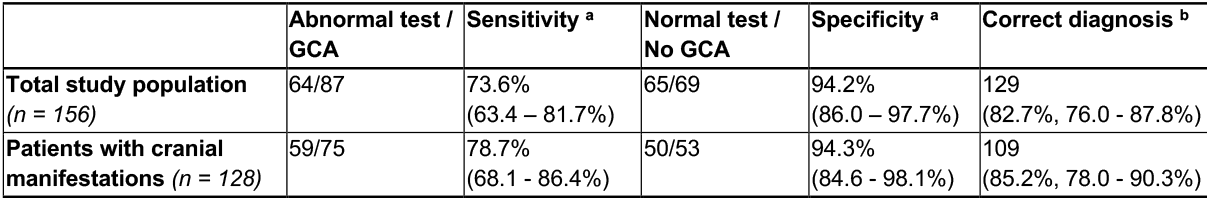
Diagnostic measures of DWI-2x compared to the reference diagnosis. ^a^, % (95%-confidence interval); ^b^, n (%, 95%-confidence interval)

|  | Abnormal test /<br>GCA | Sensitivity <sup>a</sup> | Normal test /<br>No GCA | Specificity <sup>a</sup> | Correct diagnosis <sup>b</sup> |
| --- | --- | --- | --- | --- | --- |
| **Total study population**<br>(*n* = 156) | 64/87 | 73.6%<br>(63.4 – 81.7%) | 65/69 | 94.2%<br>(86.0 – 97.7%) | 129<br>(82.7%, 76.0 - 87.8%) |
| **Patients with cranial**<br>**manifestations** (*n* = 128) | 59/75 | 78.7%<br>(68.1 - 86.4%) | 50/53 | 94.3%<br>(84.6 - 98.1%) | 109<br>(85.2%, 78.0 - 90.3%) |

The inter-rater analysis for DWI-2 and T1-BB was based on 12 patients with and 8 without GCA, including 167 and 163 segments for DWI-2 and T1-BB respectively. Compared to the reference diagnosis, the correct diagnosis was given by both readers in 19/20 (95%, 95%-CI 76.4 - 99.1%) for DWI-2 and 18/20 (90%, 95%-CI 69.9 - 97.2%) for T1-BB. The binary agreement on the patient level was 100% (n = 20, 95%-CI 83.9 - 100.0%), with a kappa of 1.00 (95%-CI not estimable) for DWI-2 and 90% (n = 18, 95%-CI 69.9 - 97.2%), with a kappa of 0.78 (95%-CI 0.50 - 1.00) for T1-BB. The binary agreement on the segment level was 93.4% (156/167, 95%-CI 88.6 - 96.3%), with a kappa of 0.85 (95%-CI 0.76 - 0.93) for DWI-2 and 89.6% (146/163, 95%-CI 83.9 - 93.4%), with a kappa of 0.79 (95%-CI 0.69 - 0.88) for T1-BB. More details on binary agreement are shown in **Supplementary Table S6**.

To determine whether the hyperintense signal in DWI images may be due to a “T2-shine through” effect, the T2-fs was directly correlated with DWI-1 (**Supplementary Table S7)**. Spearman’s rho was 0.74 (95%-CI: 0.71 – 0.76), demonstrating a strong correlation between the two scores [32].

## Discussion

Our study about the diagnostic performance of DWI for the diagnosis of GCA demonstrates a very good diagnostic accuracy, comparable to the current MRI reference sequence, the T1-BB. The sensitivity (75.9%) and specificity (94.2%) of the DWI-2 method are almost equal to the T1-BB in the largest prospective study to date (sensitivity 78.4%, specificity 90.4%) [24]. They are also in line with the pooled values for MRI of the head (T1-BB sequence) from the meta-analysis informing the 2018 EULAR recommendations for imaging in large-vessel vasculitis (sensitivity 73%, specificity 88%) [6,7]. Compared to DWI-2, the T1-BB shows a better sensitivity (88.5%) but a slightly lower specificity (88.4%) for the diagnosis of GCA in the total study population. While the diagnostic reference standard was identical to most of the recent prospective studies on the use of MRI for suspected GCA, there are some relevant differences in methodology. To reflect clinical practice as closely as possible, there were no prerequisites for signs or symptoms and patients were not excluded if CS were started *or* a biopsy was performed before imaging. Some of these exclusion criteria were used in recent prospective studies [24,33–35]. Also, the T1-BB sequence was acquired with 30 slices, whereas typically about 20 slices are acquired [21–24]. In addition, a limited number of patients with relapses were included. However, from an imaging perspective, they can be considered equivalent to new-onset GCA as all presented with new-onset cranial manifestations and showed vasculitis of the SCAs with MRI.

Interestingly, the exploratory analysis with results only from bilateral frontal and parietal branches (DWI-2x) shows only a slightly inferior diagnostic performance compared with DWI-2, whereas the time requirement for image analysis is significantly lower. While it is well known that the occipital artery is rarely affected in isolation, it is surprising that omission of both CSTAs has so little effect on diagnostic accuracy [36].

With an AUC of 0.90, DWI has a very good diagnostic performance, and the estimated best cut-off of ≥ 2 slices confirms the predefined DWI-2 as the best DWI method in suspected GCA. Clinicians are interested in knowing at what cut-off imaging allows a truly confident GCA diagnosis. With a specificity of 98.6% and a positive likelihood ratio of 48.4 for a cut-off of ≥ 3 slices with a DWI score of 2 *or* 3, an accurate diagnosis is possible even in the setting of low to moderate pre-test probability.

Due to its low specificity, the DWI-1 is not suitable for the diagnosis of GCA. This was expected from our previous experience in clinical practice and is possibly due to the low spatial resolution and imperfect anatomical correlations. A perfectly aligned slice in combination with perfect correlation of the crosshair with the 3D-TOF-MRA may have led to higher specificity, but this is not realistically achieved. DWI-1 was mainly included in the current study to be used as comparison with T2-fs on a single slice. Its high sensitivity and low negative likelihood ratio (0.06) could support decision making in cases with borderline T1-BB results.

The strength of agreement in the inter-rater analysis is perfect for DWI-2 and substantial for T1-BB for the patient level and almost perfect for DWI-2 and substantial for T1-BB for the segment level [37,38]. While the inter-rater analysis was limited to twenty patients and the 95%-CI for Cohen’s kappa for the T1-BB at the patient level was correspondingly broad, the analysis for more than 160 segments is more reliable. The observation that segment-level agreement is higher for DWI-2 than for T1-BB, underscores the practicability of the proposed new scoring method. Overall, the degree of the observed agreements for DWI-2 and T1-BB is comparable to or slightly better than in prospective studies of T1-BB imaging in which these parameters were also determined [24,33].

Because infiltration by inflammatory cells can lead to mild diffusion restriction in combination with a “T2-shine-through” effect, e.g., in orbital inflammatory syndromes, regions of inflammation are commonly more apparent on DWI images with high b-values [9,14,16]. Because it was not possible to reliably delineate SCAs in the corresponding ADC map, it is unclear if an abnormal DWI signal (score 2 or 3) of a SCA represents abnormally low ADC values, a “T2-shine-through” effect, or both. The latter seems most likely, as 96.3% of segments with perivascular oedema on T2-fs, showed a DWI-1 score ≥ 2 on the corresponding slice. The exact pathophysiology of the SCA signal detected in DWI is not relevant for its intended use in the diagnosis of GCA.

Thirty-five of 208 (16.8%) patients were excluded due to a missing T1-BB sequence. This underscores that in a substantial proportion of patients, GCA was either not part of the initial differential diagnosis or the wrong protocol was performed. In these cases, MRI was considered insufficient for the assessment of SCAs, whereas the examination of DWI images would have been valuable.

The study has the following limitations. Due to the retrospective design, there is a risk of selection bias. While this may be relevant for comparing results with other studies, it is not problematic for intraindividual comparisons between methods. Patients excluded due to a missing T1-BB sequence were predominantly non-GCA cases (24 vs. 11), most likely because an MRI is usually only repeated in patients with high pre-test probability at the study centre, which is a vasculitis referral centre. These aspects may partly explain the relatively high proportion of GCA cases in our cohort, which, however, is similar to large prospective studies for T1-BB MRI [24,33]. The proportion of correct diagnosis is strongly influenced by prevalence; it is mainly provided for comparison between methods. It is also known that many vasculitides other than GCA can rarely affect SCAs [39,40]. Two out of four excluded patients with non-GCA vasculitis had a pathological MRI of the SCAs and two had a biopsy showing vasculitis. Because MRI of the SCAs cannot distinguish between different vasculitides, and involvement of the SCAs is extremely rare in non-GCA vasculitides, it would have been incorrect to classify these cases as false-positive or false-negative. The exclusion of 4/208 (1.9%) patients most likely did not have any relevant impact on the results. A mixture of several scanner manufacturers would have been desirable to demonstrate feasibility across platforms. However, evaluation of images from four different 3-Tesla scanner models with different DWI sequences over a four-year period shows that the DWI-scoring is applicable in a setting of varying 3-Tesla scanners. Whether analysis of the DWI sequence from 1.5-Tesla scanners would yield similar results cannot be answered by our study. Prospective studies are needed to determine the definitive value of DWI for the diagnosis of GCA, especially to reliably inform on measures dependent on prevalence.

Due to its lower spatial resolution, DWI is not expected to have a higher sensitivity than the T1-BB sequence but showed a higher specificity and an excellent inter-rater agreement, features important for the diagnosis of GCA. The DWI signal of SCAs with vasculitis disappears with therapy and returns at relapse, which may even allow its use for follow-up studies. Analysis of the DWI sequence may also be useful as a case-finding strategy in patients older than 50 years who undergo an MRI scan for headache or visual disturbances, even if GCA is not initially suspected.

The DWI sequence does not require the application of contrast agents, has a short acquisition time, and is exceptionally widely available. Together with the good sensitivity and excellent specificity for the diagnosis of GCA, the proposed DWI rating method for SCAs represents a valuable alternative to the T1-BB sequence, especially in emergency or unclear clinical situations. Because DWI is already part of most protocols for MRI of the head, it can be applied immediately in clinical practice.

## Key Messages

- DWI can identify GCA of the superficial cranial arteries (sensitivity: 75.9%, specificity: 94.2%).
- A pathological DWI-signal over ≥ 3 slices has a positive LR of 48.4 for GCA diagnosis.
- DWI is available worldwide and can be used immediately in clinical practice for suspected GCA.

## Supporting information

Supplementary Atlas

## Contributors

**LS** initiated and led the project, obtained ethical approval, identified patients, programmed the database, co-determined the reference diagnosis, defined the DWI- and T2-fs-scores, re-read MR images, wrote the statistical analysis plan, analysed, and interpreted the data, and wrote the manuscript. **SB** identified patients, compiled the clinical data and sequence parameters, supported the set-up of the database, contributed to the statistical analysis plan, analysed, and interpreted the data, created images for the atlas, compiled the tables, and co-wrote the manuscript. **LB** reviewed the statistical analysis plan, performed the statistical analysis, and contributed to the manuscript. **BM** assisted in obtaining ethical approval, contributed to data analysis and interpretation, and revised the manuscript. **HMB** supported the project as a radiologist with long-term experience in imaging of vasculitis, contributed to the data analysis, and revised the manuscript. **FW** assisted in obtaining ethical approval, supported the data analysis and revised the manuscript. **FL** co-defined DWI- and T2-fs-scores, identified patients, co-determined the reference diagnosis, assisted in obtaining ethical approval, analysed and interpreted the data, and co-wrote the manuscript. **PS** co-defined the DWI- and the T2-fs-scores, identified patients, co-determined the reference diagnosis, co-wrote the statistical analysis plan, re-read MR images, compiled the supplementary atlas, analysed, and interpreted the data, and co-wrote the manuscript.

## Data availability

The data underlying this article will be shared on reasonable request to the corresponding author.

