## Supplementary Material for "Diffusion-weighted magnetic resonance imaging for the diagnosis of giant cell arteritis – a comparison with T1-weighted black-blood imaging"

Supplementary Figures S1 – S4

Supplementary Tables S1 – S7

**Supplementary Figure S1. Patient flow chart**

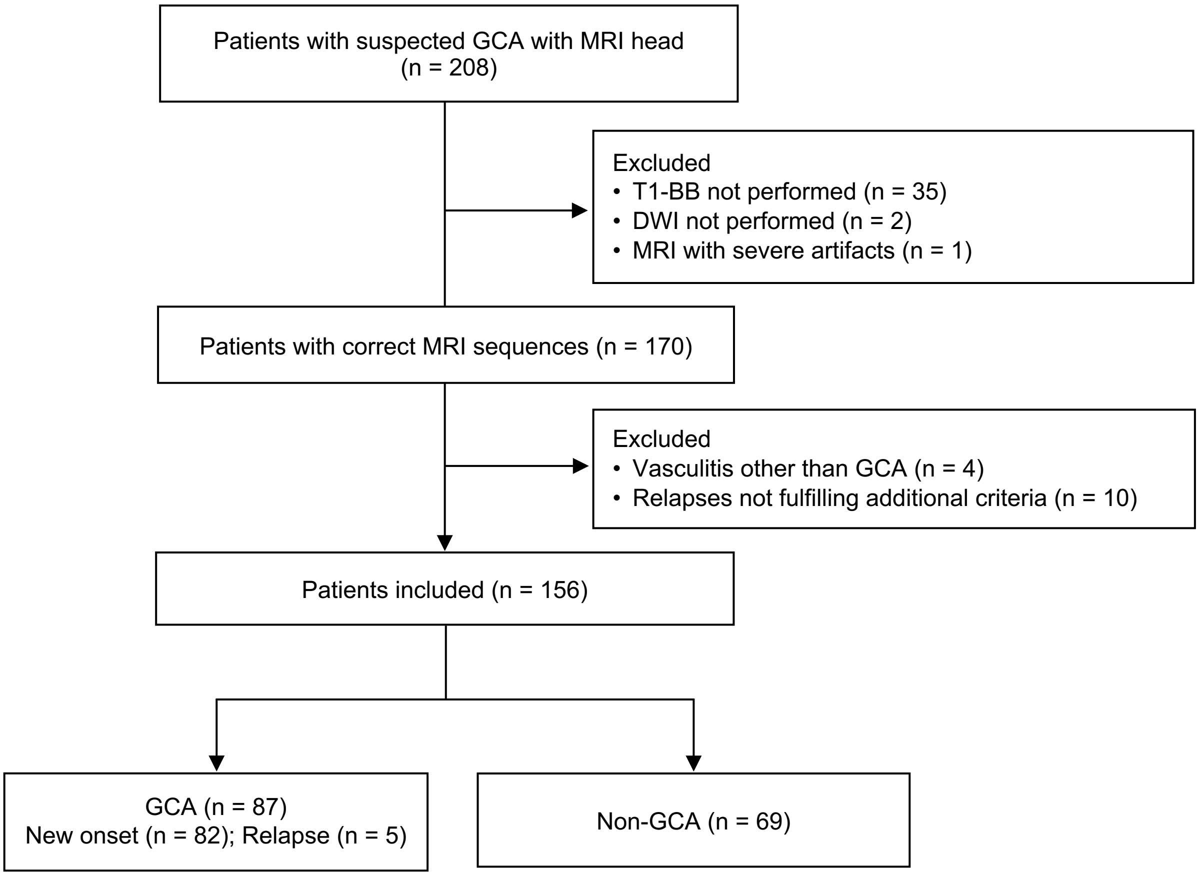


**Figure legend S1**: Vasculitis other than GCA: 1 x ANCA-associated vasculitis, 1x IgG4-related temporal arteritis, 1x drug-induced-vasculitis, 1x transient and self-limited vasculitis of unknown cause.

### Supplementary Table S1. Details of MRI scanners, DWI sequences and parameters

| Characteristics | N = 156 (n, %) |
|---|---|
| **MRI scanner** | |
| Siemens Prisma Fit | 15 (9.6%) |
| Siemens Skyra Fit | 40 (25.6%) |
| Siemens Verio | 2 (1.3%) |
| Siemens Vida | 99 (63.5%) |
| **Receiving coil** | |
| HeadNeck 20 channels | 37 (23.7%) |
| HeadNeck 64 channels | 61 (39.1%) |
| Unknown | 58 (37.2%) |
| **DWI method** | |
| EPI, readout-segmented multi-shot (RESOLVE) | 154 (98.7%) |
| EPI, single shot | 2 (1.3%) |
| **b-value** | |
| 1000 s/mm$^2$ | 156 (100%) |
| **Slice thickness** | |
| 4 mm | 154 (98.7%) |
| 5 mm | 2 (1.3%) |
| **Repetition time (TR) (ms)** | |
| Median (IQR) | 5750 (4570, 5750) |
| Range | 3500, 5750 |
| **Echo time (TE) (ms)** | |
| Median (IQR) | 61 (61, 67) |
| Range | 55, 89 |
| **Number of excitations / averages** | |
| 1 | 154 (98.7%) |
| 4 | 2 (1.3%) |
| **Acquisition matrix** | |
| 128 x 128 | 2 (1.3%) |
| 176 x 176 | 8 (5.1%) |
| 192 x 192 | 146 (93.6%) |
| **Field of view** | |
| 220 mm x 220 mm | 154 (98.7%) |
| 230 mm x 230 mm | 2 (1.3%) |
| **Voxel size** | |
| 1.15 x 1.15 x 4.00 mm | 146 (93.6%) |
| 1.25 x 1.25 x 4.00 mm | 8 (5.1%) |
| 1.80 x 1.80 x 5.00 mm | 2 (1.3%) |
| **Flip angle** | |
| 90 | 2 (1.3%) |
| 180 | 154 (98.7%) |
| **Slice spacing** | |
| 4.8 mm | 153 (98.1%) |
| 5.2 mm | 1 (0.6%) |
| 6.0 mm | 2 (1.3%) |

**Table legend S1:** EPI, echo-planar imaging; IQR, interquartile range

**Supplementary Figure S2. Brightness setting for assessment of superficial cranial arteries in DWI**

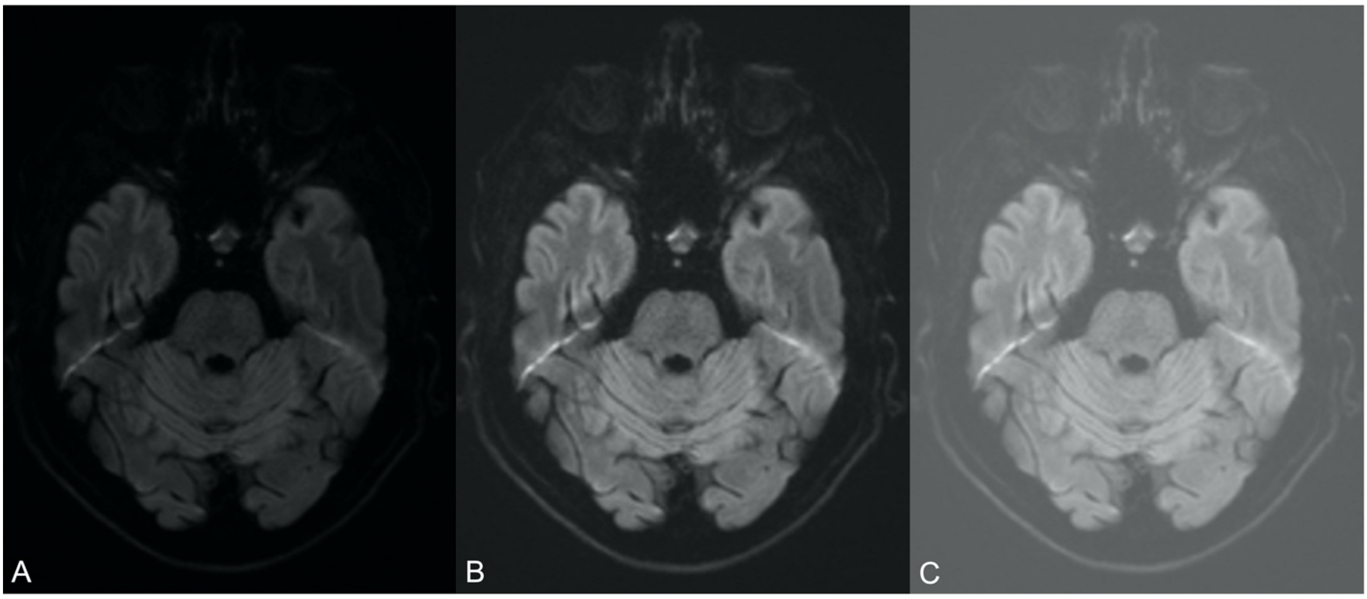


**Figure legend S2: A**) The brightness is too low: skin and air are not distinguishable; the subcutaneous fat is black. **B**) Optimal brightness: the skin can be distinguished from the surrounding air; the subcutaneous fat is dark gray and anatomical structures such as facial muscles are readily visible. **C**) Brightness is too high.

**Supplementary Figure S3. T2-fs-scoring for superficial cranial arteries**

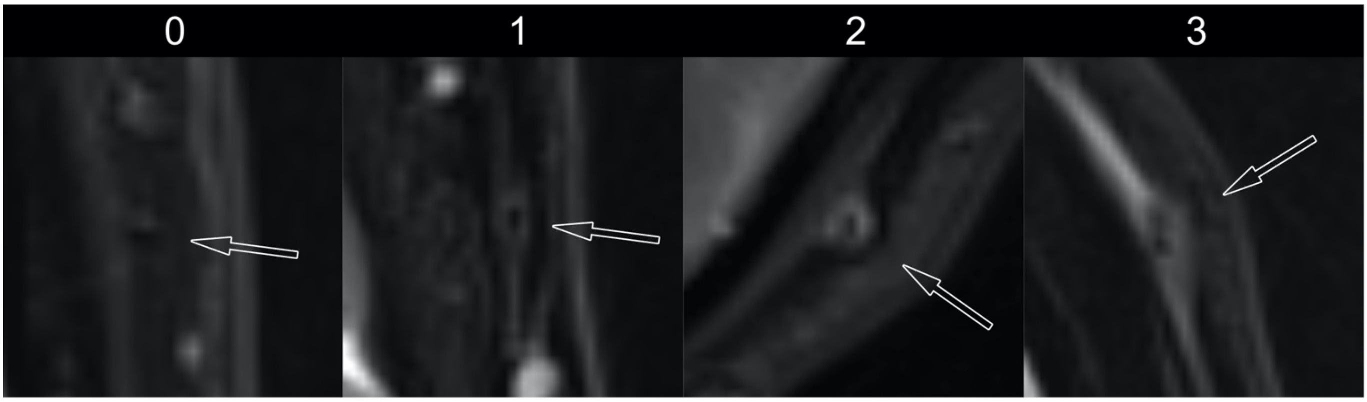


**Figure legend S3:** T2-fs-scoring (0 – 3): 0 = vessel wall not visible; 1= vessel wall visible; 2 = vessel wall prominently visible; 3 = vessel wall prominently visible and perivascular edema.

**Supplementary Figure S4. DWI-scoring for superficial cranial arteries (version without arrows)**

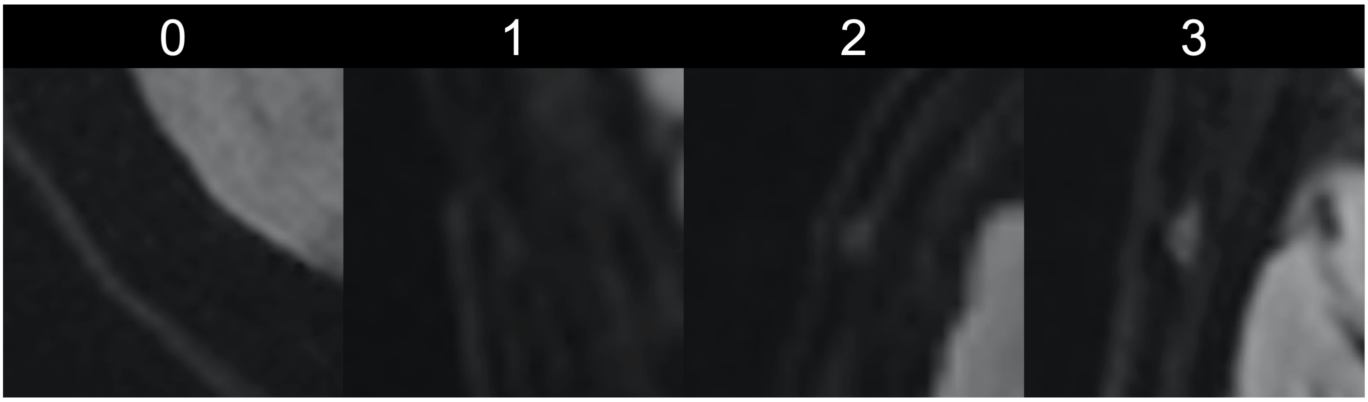


**Figure legend S4:** DWI-scoring (0 – 3): 0 = artery not visible; 1 = artery slightly visible; 2 = artery prominently visible; 3 = artery brightly visible.

**Supplementary Table S2. Clinical diagnosis for patients without giant cell arteritis (total study population)**

| Expert Diagnosis at 6 months | n (%) *(N = 69)* |
|---|---|
| Polymyalgia rheumatica | 23 (33.3%) |
| Primary headache | 10 (14.5%) |
| Non-arteritic anterior ischemic optic neuropathy | 9 (13.0%) |
| Polyarthritis | 9 (13.0%) |
| Retinal artery occlusion | 2 (2.9%) |
| Prominently visible temporal artery (physiological) | 2 (2.9%) |
| Infection | 2 (2.9%) |
| Lymphoma | 2 (2.9%) |
| Sarcoidosis | 2 (2.9%) |
| Other * | 8 (11.6%) |

* One each: atherosclerosis; calcium pyrophosphate deposition disease (CPPD); cryptogenic organizing pneumonia; hypertensive urgency; idiopathic abducens palsy; late-onset spondyloarthritis; stroke; unclear systemic inflammation with spontaneous resolution

**Supplementary Table S3. Correct diagnosis of DWI-1, DWI-2 and T1-BB methods compared to the reference diagnosis**

**Total study population ***

| | Total *(N = 156)* | No GCA *(N = 69)* | GCA *(N = 87)* | Correct diagnosis *(N = 156)* |
|---|---|---|---|---|
| **DWI-1** | | | | 111 (71.2%, 63.6 - 77.7%) |
| No Vasculitis | 28 (17.9%, 12.7 - 24.7%) | 26 (37.7%, 27.2 - 49.5%) | 2 (2.3%, 0.6 - 8.0%) | |
| Vasculitis | 128 (82.1%, 75.3 - 87.3%) | 43 (62.3%, 50.5 - 72.8%) | 85 (97.7%, 92.0 - 99.4%) | |
| **DWI-2** | | | | 131 (84.0%, 77.4 - 88.9%) |
| No Vasculitis | 86 (55.1%, 47.3 - 62.7%) | 65 (94.2%, 86.0 - 97.7%) | 21 (24.1%, 16.4 - 34.1%) | |
| Vasculitis | 70 (44.9%, 37.3 - 52.7%) | 4 (5.8%, 2.3 - 14.0%) | 66 (75.9%, 65.9 - 83.6%) | |
| **T1-BB** | | | | 138 (88.5%, 82.5 - 92.6%) |
| No Vasculitis | 71 (45.5%, 37.9 - 53.3%) | 61 (88.4%, 78.8 - 94.0%) | 10 (11.5%, 6.4 - 19.9%) | |
| Vasculitis | 85 (54.5%, 46.7 - 62.1%) | 8 (11.6%, 6.0 - 21.2%) | 77 (88.5%, 80.1 - 93.6%) | |

**Patients with cranial manifestations ***

| | Total *(N = 128)* | No GCA *(N = 53)* | GCA *(N = 75)* | Correct diagnosis *(N = 128)* |
|---|---|---|---|---|
| **DWI-1** | | | | 92 (71.9%, 63.5 - 78.9%) |
| No Vasculitis | 21 (16.4%, 11.0 - 23.8%) | 19 (35.8%, 24.3 - 49.3%) | 2 (2.7%, 0.7 - 9.2%) | |
| Vasculitis | 107 (83.6%, 76.2 - 89.0%) | 34 (64.2%, 50.7 - 75.7%) | 73 (97.3%, 90.8 - 99.3%) | |
| **DWI-2** | | | | 111 (86.7%, 79.8 - 91.5%) |
| No Vasculitis | 64 (50.0%, 41.5 - 58.5%) | 50 (94.3%, 84.6 - 98.1%) | 14 (18.7%, 11.5 - 28.9%) | |
| Vasculitis | 64 (50.0%, 41.5 - 58.5%) | 3 (5.7%, 1.9 - 15.4%) | 61 (81.3%, 71.1 - 88.5%) | |
| **T1-BB** | | | | 118 (92.2%, 86.2 - 95.7%) |
| No Vasculitis | 53 (41.4%, 33.2 - 50.1%) | 48 (90.6%, 79.7 - 95.9%) | 5 (6.7%, 2.9 - 14.7%) | |
| Vasculitis | 75 (58.6%, 49.9 - 66.8%) | 5 (9.4%, 4.1 - 20.3%) | 70 (93.3%, 85.3 - 97.1%) | |

**Table legend S3:** *, n (%, 95%-Confidence interval); DWI-1, diffusion weighted imaging with ≥ 1 slice; DWI-2, diffusion weighted imaging with ≥ 2 slices; T1-BB, T1-black-blood.

**Supplementary Table S4. Comparison of DWI-2 with T1-BB on a segment level (total study population)**

| | *N** | **Total** *n (%)* | *N** | **No GCA** *n (%)* | *N** | **GCA** *n (%)* | **P-value** |
|---|---|---|---|---|---|---|---|
| **Common superficial temporal arteries** | 254 | | 121 | | 133 | | <0.001 |
| DWI-2 and T1-BB normal | | 173 (68.1%) | | 117 (96.7%) | | 56 (42.1%) | |
| DWI-2 pathological, T1-BB normal | | 1 (0.4%) | | 0 (0.0%) | | 1 (0.8%) | |
| DWI-2 normal, T1-BB pathological | | 32 (12.6%) | | 3 (2.5%) | | 29 (21.8%) | |
| DWI-2 and T1-BB pathological | | 48 (18.9%) | | 1 (0.8%) | | 47 (35.3%) | |
| **Temporal arteries, frontal branches** | 309 | | 137 | | 172 | | <0.001 |
| DWI-2 and T1-BB normal | | 173 (56.0%) | | 128 (93.4%) | | 45 (26.2%) | |
| DWI-2 pathological, T1-BB normal | | 2 (0.6%) | | 2 (1.5%) | | 0 (0.0%) | |
| DWI-2 normal, T1-BB pathological | | 27 (8.7%) | | 4 (2.9%) | | 23 (13.4%) | |
| DWI-2 and T1-BB pathological | | 107 (34.6%) | | 3 (2.2%) | | 104 (60.5%) | |
| **Temporal arteries, parietal branches** | 295 | | 134 | | 161 | | <0.001 |
| DWI-2 and T1-BB normal | | 198 (67.1%) | | 131 (97.8%) | | 67 (41.6%) | |
| DWI-2 pathological, T1-BB normal | | 3 (1.0%) | | 0 (0.0%) | | 3 (1.9%) | |
| DWI-2 normal, T1-BB pathological | | 27 (9.2%) | | 3 (2.2%) | | 24 (14.9%) | |
| DWI-2 and T1-BB pathological | | 67 (22.7%) | | 0 (0.0%) | | 67 (41.6%) | |
| **Posterior auricular arteries** | 78 | | 41 | | 37 | | <0.001 |
| DWI-2 and T1-BB normal | | 62 (79.5%) | | 40 (97.6%) | | 22 (59.5%) | |
| DWI-2 pathological, T1-BB normal | | 0 (0.0%) | | 0 (0.0%) | | 0 (0.0%) | |
| DWI-2 normal, T1-BB pathological | | 12 (15.4%) | | 1 (2.4%) | | 11 (29.7%) | |
| DWI-2 and T1-BB pathological | | 4 (5.1%) | | 0 (0.0%) | | 4 (10.8%) | |
| **Occipital arteries** | 308 | | 136 | | 172 | | <0.001 |
| DWI-2 and T1-BB normal | | 199 (64.6%) | | 131 (96.3%) | | 68 (39.5%) | |
| DWI-2 pathological, T1-BB normal | | 2 (0.6%) | | 0 (0.0%) | | 2 (1.2%) | |
| DWI-2 normal, T1-BB pathological | | 31 (10.1%) | | 2 (1.5%) | | 29 (16.9%) | |
| DWI-2 and T1-BB pathological | | 76 (24.7%) | | 3 (2.2%) | | 73 (42.4%) | |
| **All segments combined** | 1244 | | 569 | | 675 | | <0.001 |
| DWI-2 and T1-BB normal | | 805 (64.7%) | | 547 (96.1%) | | 258 (38.2%) | |
| DWI-2 pathological, T1-BB normal | | 8 (0.6%) | | 2 (0.4%) | | 6 (0.9%) | |
| DWI-2 normal, T1-BB pathological | | 129 (10.4%) | | 13 (2.3%) | | 116 (17.2%) | |
| DWI-2 and T1-BB pathological | | 302 (24.3%) | | 7 (1.2%) | | 295 (43.7%) | |

**Table legend S4:** * Number of non-missing observations; DWI-2, diffusion weighted imaging with ≥ 2 slices; T1-BB, T1-black-blood.

**Supplementary Table S5. Sensitivity and specificity of DWI-2 with T1-BB as reference (total study population)**

| | **Pathological DWI-2 / Pathological T1-BB** | **Sensitivity (95% CI)** | **Normal DWI-2 / Normal T1-BB** | **Specificity (95% CI)** |
|---|---|---|---|---|
| Left common superficial TA | 20/38 | 52.6% (37.3 - 67.5%) | 86/86 | 100.0% (95.7 - 100.0%) |
| Right common superficial TA | 28/42 | 66.7% (51.6 - 79.0%) | 87/88 | 98.9% (93.8 - 99.8%) |
| Left TA frontal branch | 49/62 | 79.0% (67.4 - 87.3%) | 90/91 | 98.9% (94.0 - 99.8%) |
| Right TA frontal branch | 58/72 | 80.6% (70.0 - 88.0%) | 83/84 | 98.8% (93.6 - 99.8%) |
| Left TA parietal branch | 33/43 | 76.7% (62.3 - 86.8%) | 100/102 | 98.0% (93.1 - 99.5%) |
| Right TA parietal branch | 34/51 | 66.7% (53.0 - 78.0%) | 98/99 | 99.0% (94.5 - 99.8%) |
| Left post. auricular artery | 1/7 | 14.3% (2.6 - 51.3%) | 29/29 | 100.0% (88.3 - 100.0%) |
| Right post. auricular artery | 3/9 | 33.3% (12.1 - 64.6%) | 33/33 | 100.0% (89.6 - 100.0%) |
| Left occipital artery | 36/50 | 72.0% (58.3 - 82.5%) | 103/103 | 100.0% (96.4 - 100.0%) |
| Right occipital artery | 40/57 | 70.2% (57.3 to 80.5%) | 96/98 | 98.0% (92.9 - 99.4%) |

**Table legend S5:** DWI-2, diffusion weighted imaging with ≥ 2 slices; T1-BB, T1-black-blood; TA, temporal artery.

### Supplementary Table S6. Binary agreement between readers for DWI-2 and T1-BB for a subset of 20 patients

| | **Reader 2 – No Vasculitis (Score 0 or 1)** n (%, 95% CI) | **Reader 2 – Vasculitis (Score 2 or 3)** n (%, 95% CI) | **Agreement** n (%, 95% CI) | **Cohen's kappa** (95% CI) |
|---|---|---|---|---|
| **Patient level (Reader 1)** | | | | |
| **DWI-2** | | | 20 (100%, 83.9 - 100%) | 1.00 (not estimable) |
| No Vasculitis | 9 (100%, 70.1 - 100%) | 0 (0.0%, 0.0 - 25.9%) | | |
| Vasculitis | 0 (0.0%, 0.0 - 29.9%) | 11 (100%, 74.1 - 100%) | | |
| **T1-BB** | | | 18 (90.0%, 69.9 - 97.2%) | 0.78 (0.50 - 1.00) |
| No Vasculitis | 6 (75.0%, 40.9 - 92.9%) | 0 (0.0%, 0.0 - 24.2%) | | |
| Vasculitis | 2 (25.0%, 7.1 - 59.1%) | 12 (100%, 75.8 - 100%) | | |
| **Segment level (Reader 1)** | | | | |
| **DWI-2 (all segments)** | | | 156 (93.4%, 88.6 - 96.3%) | 0.85 (0.76 - 0.93) |
| Score 0 or 1 | 108 (95.6%, 90.1 - 98.1%) | 6 (11.1%, 5.2 - 22.2%) | | |
| Score 2 or 3 | 5 (4.4%, 1.9 - 9.9%) | 48 (88.9%, 77.8 - 94.8%) | | |
| **T1-BB (all segments)** | | | 146 (89.6%, 83.9 - 93.4%) | 0.79 (0.69 - 0.88) |
| Score 0 or 1 | 87 (87.9%, 80.0 - 92.9%) | 5 (7.8%, 3.4 - 17.0%) | | |
| Score 2 or 3 | 12 (12.1%, 7.1 - 20.0%) | 59 (92.2%, 83.0 - 96.6%) | | |

**Table legend S6:** CI, confidence interval; DWI-2, diffusion weighted imaging with ≥ 2 slices; T1-BB, T1-black-blood.

### Supplementary Table S7. Correlation of DWI-1 and T2-fs on a segment level

| | **T2-fs score** | | | |
|---|---|---|---|---|
| | **0** *(N = 77)* | **1** *(N = 663)* | **2** *(N = 279)* | **3** *(N = 220)* |
| **DWI-1 score** - n (%) | | | | |
| **0** | 48 (62.3%) | 64 (9.7%) | 0 (0.0%) | 0 (0.0%) |
| **1** | 27 (35.1%) | 503 (75.9%) | 99 (35.5%) | 4 (1.8%) |
| **2** | 1 (1.3%) | 95 (14.3%) | 160 (57.3%) | 105 (47.7%) |
| **3** | 0 (0.0%) | 1 (0.2%) | 17 (6.1%) | 107 (48.6%) |
| **missing** | 1 (1.3%) | 0 (0.0%) | 3 (1.1%) | 4 (1.8%) |

**Table legend S7:** Based on 1239 segments from 152 patients (T2-fs was missing for 4 patients). Spearman's rho: 0.74 (95%-confidence interval: 0.71 – 0.76). DWI-1, diffusion weighted imaging with ≥ 1 slice.
