## Supplementary Atlas for "Diffusion-weighted magnetic resonance imaging for the diagnosis of giant cell arteritis – a comparison with T1-weighted black-blood imaging"

The available literature and example images on diffusion-weighted imaging (DWI) for the diagnosis of giant cell arteritis are currently limited. The images and accompanying information in this atlas are intended to assist the reader in successfully using DWI for the diagnosis of giant cell arteritis in daily clinical practice. Corresponding T1-black-blood (T1-BB) or 3D arterial time-of-flight MR angiography (3D-TOF-MRA) images are provided for comparison.

Slightly different anatomical positions of the corresponding slices are unavoidable due to a slice thickness of 4 - 5 mm for DWI and 3 mm for T1-BB (3mm gap between slices) and the slightly different orientation of the axial plane in different sequences.

All images in this atlas were acquired in the axial plane. DWI images are shown with a b-value of 1000 s/mm$^2$.

The black background of the atlas was chosen to reflect the setting of image analysis in clinical practice.

---

**Atlas Table 1** Illustrates corresponding anatomy of axial DWI and T1-BB images on three different levels: below the external ear canal, at the midlevel of the orbit, above the external ear canal.

**Atlas Table 2** Shows examples for the DWI scores of superficial cranial arteries.

**Atlas Table 3** Shows examples for DWI of the vertebral arteries with vasculitis.

**Atlas Table 4** Demonstrates pitfalls of DWI assessment of superficial cranial arteries.

**Atlas Table 5** Outlines pitfalls in T1-BB imaging of superficial cranial arteries and indicates in which situations DWI may be helpful.

**Atlas Table 6** Shows the change in DWI scores during follow-up and relapse of giant cell arteritis.

---

#### Abbreviations

| | |
|---|---|
| 3D-TOF-MRA | 3D arterial time-of-flight MR angiography |
| CSTA | common superficial temporal artery |
| DWI | diffusion-weighted imaging |
| T1-BB | T1-black-blood |

### Atlas Table 1. Anatomy in DWI and corresponding T1-BB images

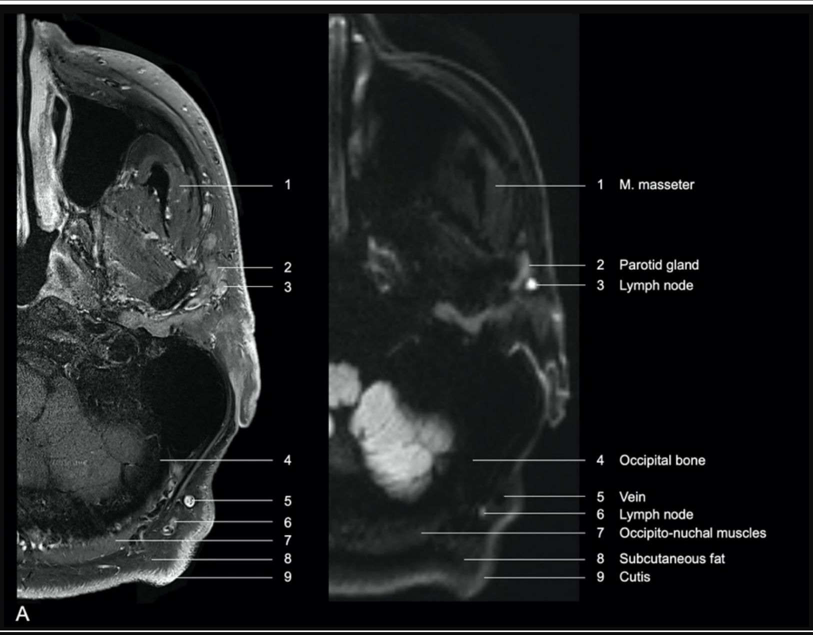


#### Axial slice below the external ear canal

The upper margin of the parotid gland can be delineated with DWI.

Normal lymph nodes are bright and well demarcated on DWI without adjacent fat signal alteration.

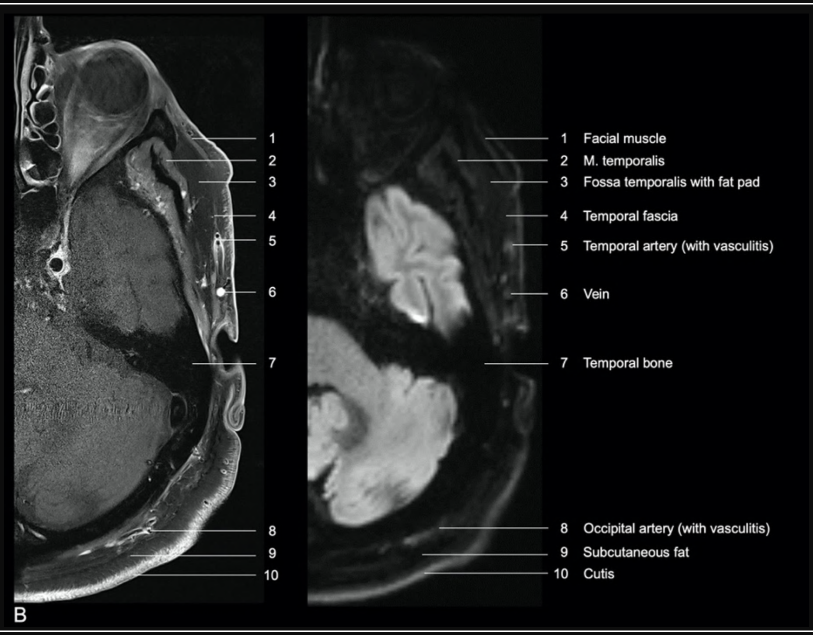


#### Axial slice at the midlevel of the orbit

The temporal artery runs superficially from the temporal fascia.

Facial muscles show a weak signal with DWI. Care must be taken not to misinterpret them as arteries. Correlation with an arterial TOF-MRA usually resolves this issue.

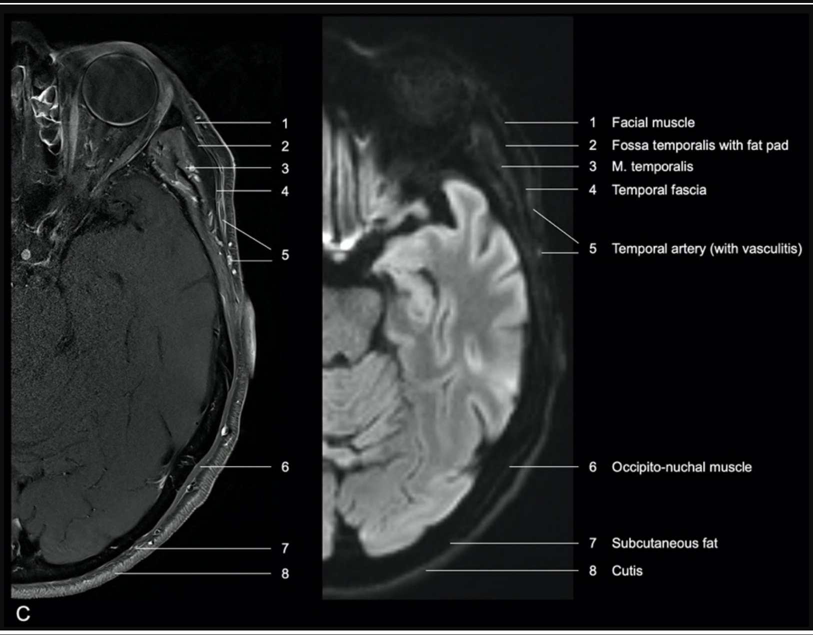


#### Axial slice above the external ear canal

The branches of the temporal artery run superficially to the temporal fascia.

The occipito-nuchal muscles are usually seen as a slightly hyperintense linear structure on DWI images at this level. The occipital artery typically runs superficial to these muscles and is sometimes difficult to distinguish.

**Atlas Table 2. Examples for DWI scoring of superficial cranial arteries**

| | | |
|---|---|---|
| **Score 1** | | |
| 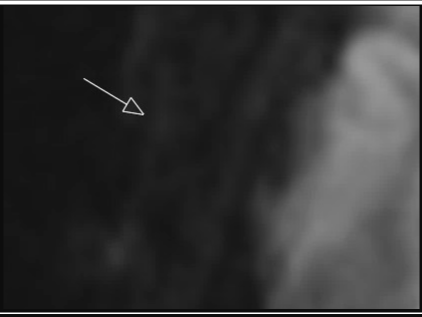 | 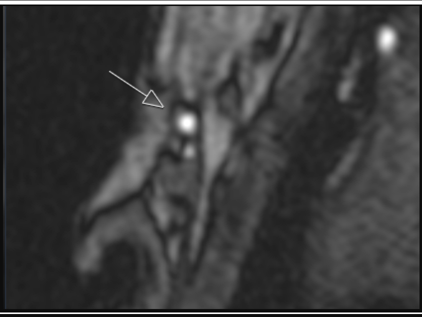 | 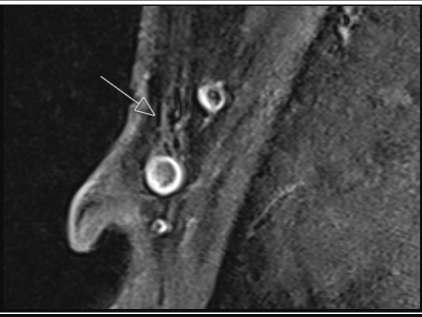 |
| DWI (right CSTA) | 3D-TOF-MRA (right CSTA) | T1-BB (right CSTA, score 1) |
| On the T1-BB image, two veins with a dark lumen could be misinterpreted as vasculitis. With the 3D-TOF-MRA, the artery can be clearly identified between the two veins. | | |
| 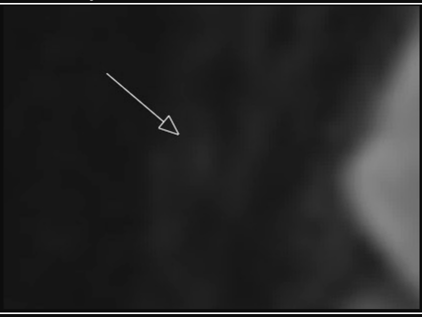 | 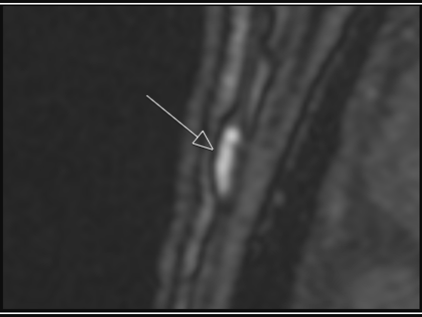 | 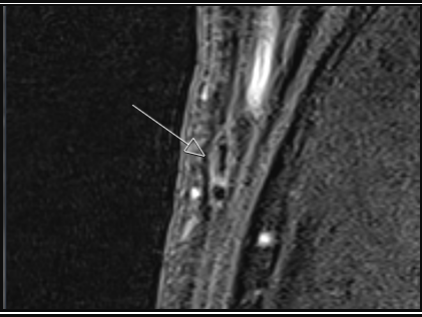 |
| DWI (right CSTA) | 3D-TOF-MRA (right CSTA) | T1-BB (right CSTA, score 1) |
| 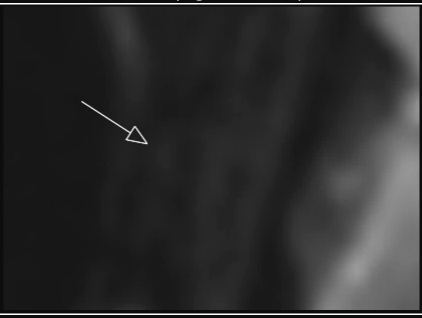 | 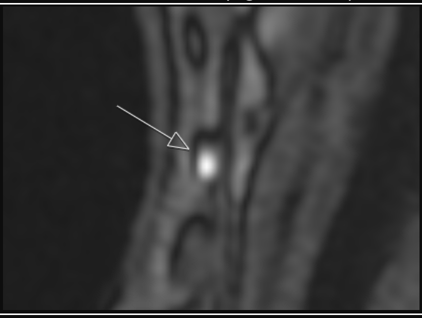 | 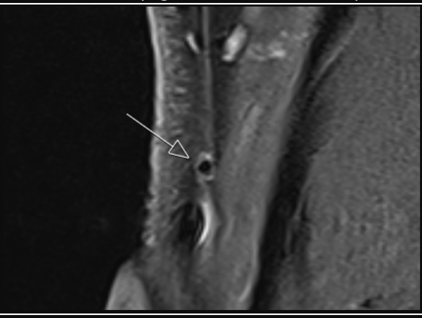 |
| DWI (right CSTA) | 3D-TOF-MRA (right CSTA) | T1-BB (right CSTA, score 1) |
| **Score 2** | | |
| 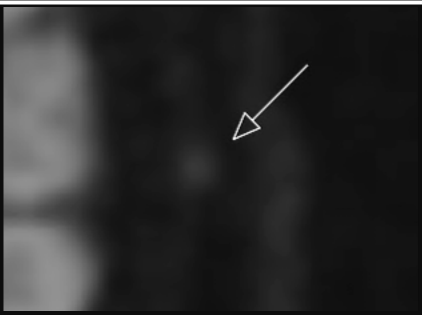 | 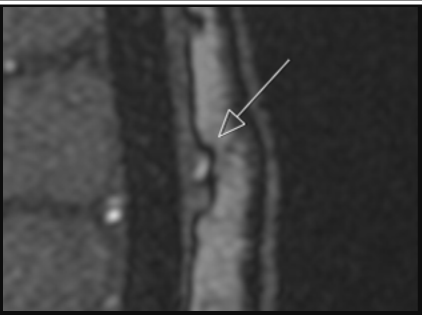 | 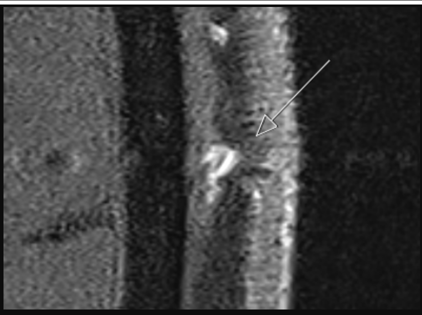 |
| DWI (left frontal branch) | 3D-TOF-MRA (left frontal branch) | T1-BB (left frontal branch, score 2) |
| 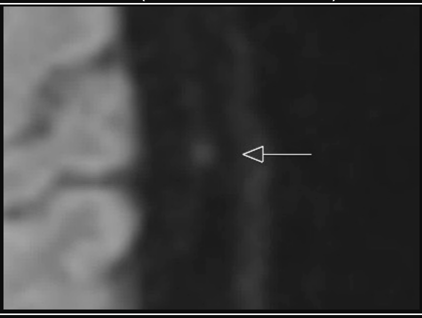 | 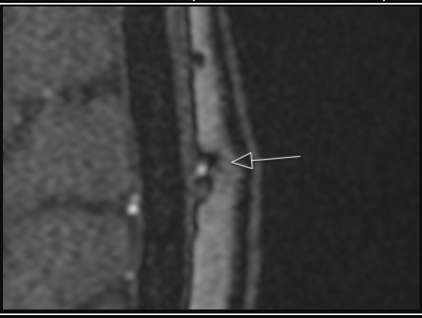 | 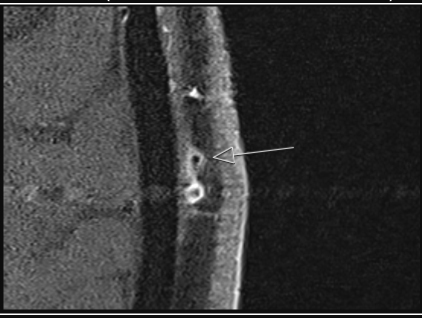 |
| DWI (left parietal branch) | 3D-TOF-MRA (left parietal branch) | T1-BB (left parietal branch, score 2) |

| | | |
|---|---|---|
| 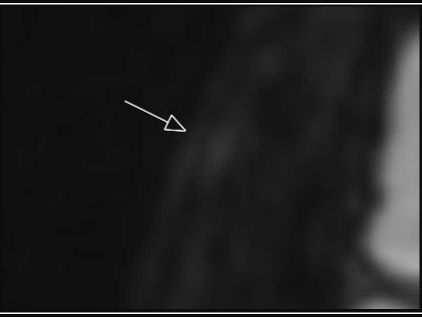 | 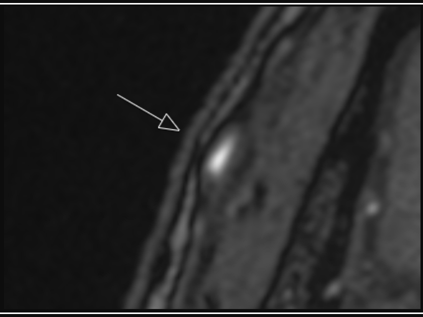 | 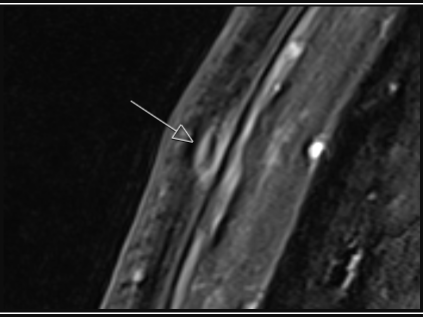 |
| DWI (right frontal branch, score 2) | 3D-TOF-MRA (right frontal branch) | T1-BB (right frontal branch, score 2) |
| **Score 3** | | |
| 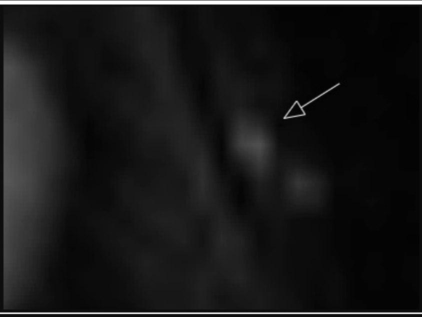 | 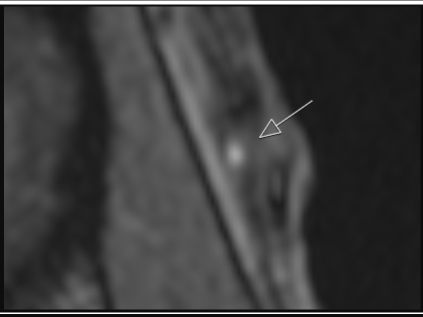 | 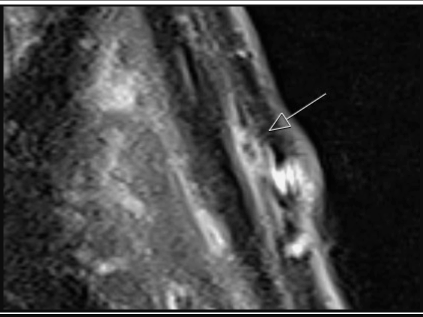 |
| DWI (left frontal branch) | 3D-TOF-MRA (left frontal branch) | T1-BB (left frontal branch, score 2) |
| 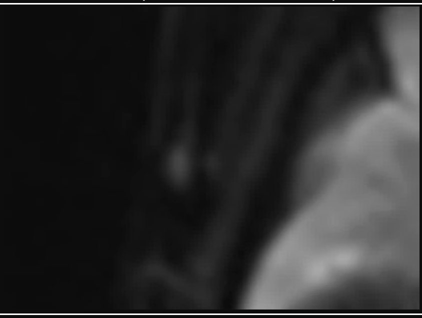 | 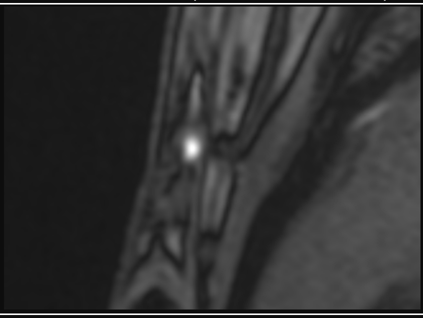 | 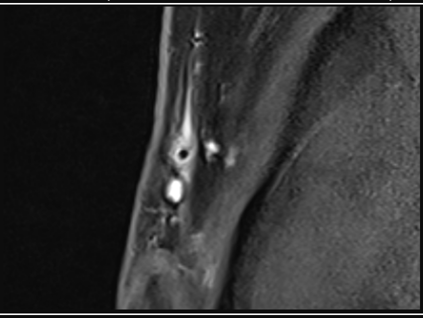 |
| DWI (right CSTA) | 3D-TOF-MRA (right CSTA) | T1-BB (right CSTA, score 3) |
| 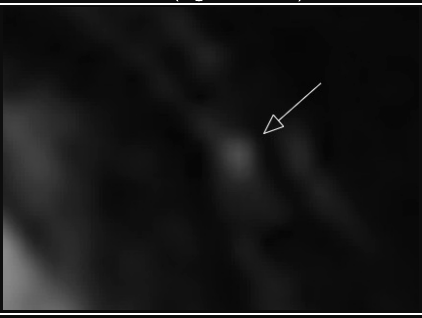 | 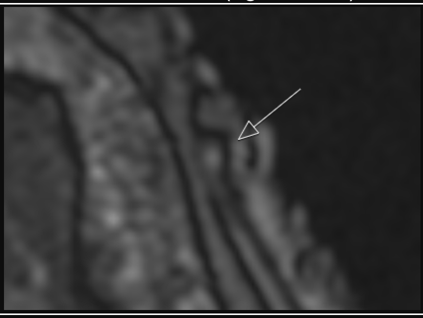 | 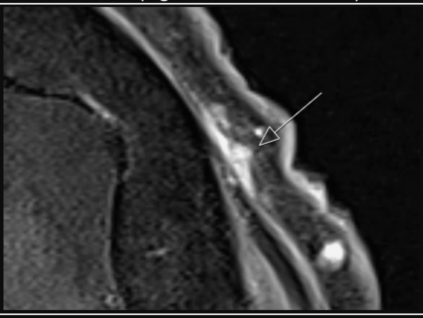 |
| DWI (left frontal branch) | 3D-TOF-MRA (left frontal branch) | T1-BB (left frontal branch, score 3) |
| **Arterial segments running in the axial plane** | | |
| 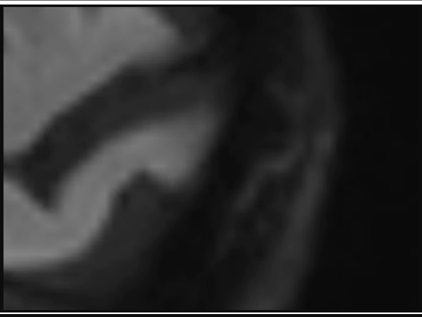 | 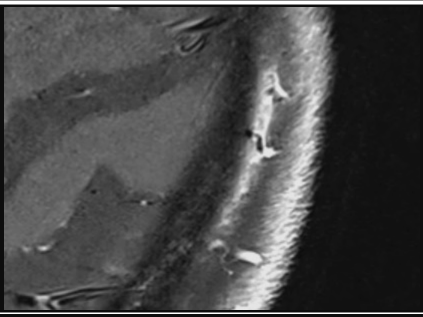 | |
| DWI (left parietal branch, score 2) | T1-BB (left parietal branch, score 3) | |

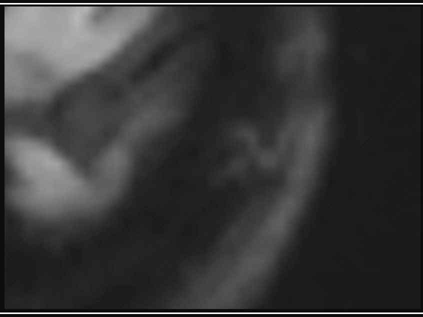
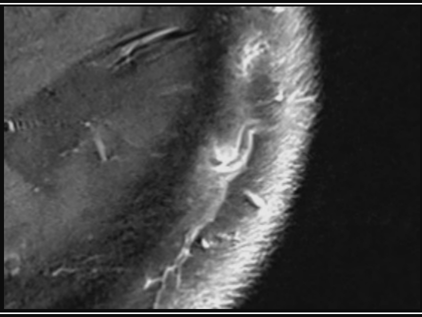

DWI (left parietal branch, score 2) T1-BB (left parietal branch, score 3)

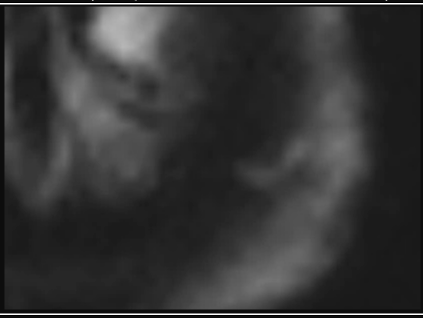
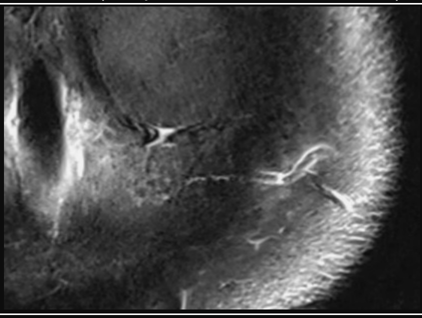

DWI (left parietal branch, score 2) T1-BB (left parietal branch, score 2)

The 3 x 2 images above show the distal left parietal branch with consecutive slices each further cranially. The vessel can be seen running in the axial plane over several millimeters in both DWI and T1-BB images. In the top image, no lumen is seen in the T1-BB due to a tangentially cut vessel and likely partial volume effects.

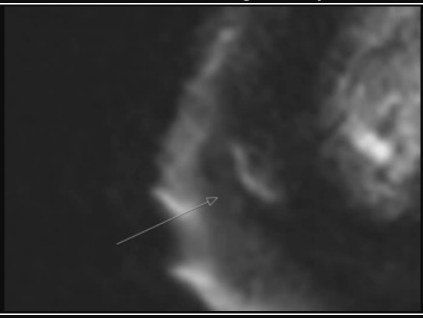
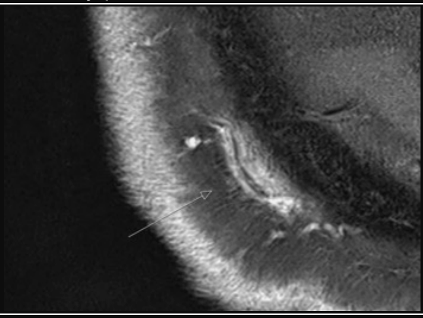

DWI (right parietal branch, score 3) T1-BB (right parietal branch, score 3)

These slices are located close to the vertex.

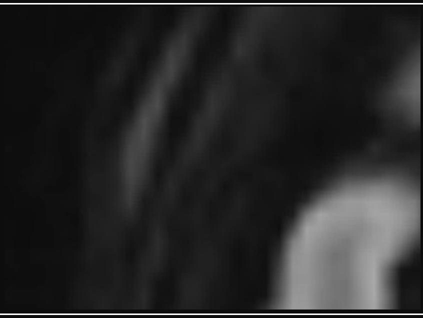
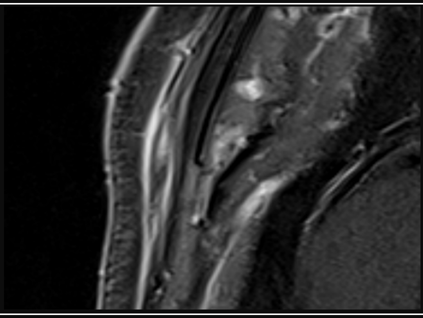

DWI (right frontal branch, score 3) T1-BB (right frontal branch, score 3)

**Very large superficial cranial arteries**

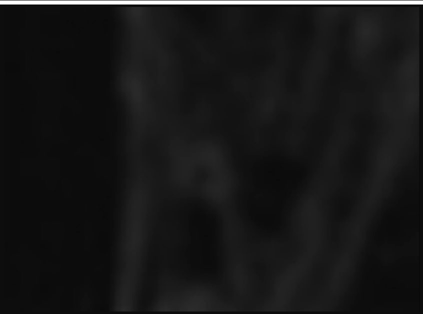
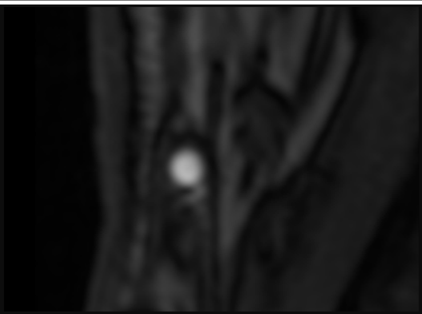
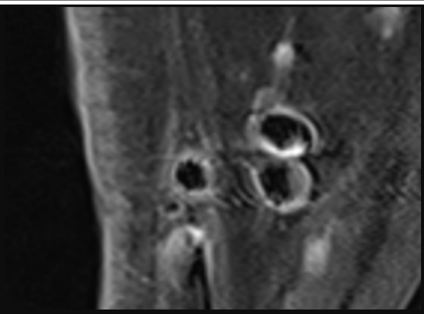

DWI (right CSTA, score 1) 3D-TOF-MRA (right CSTA) T1-BB (right CSTA, score 1)

In SCAs with very large diameters, mostly the CSTA, the lumen and vessel walls can sometimes be visualized individually. In these cases, the signal intensity of a single wall is rated.

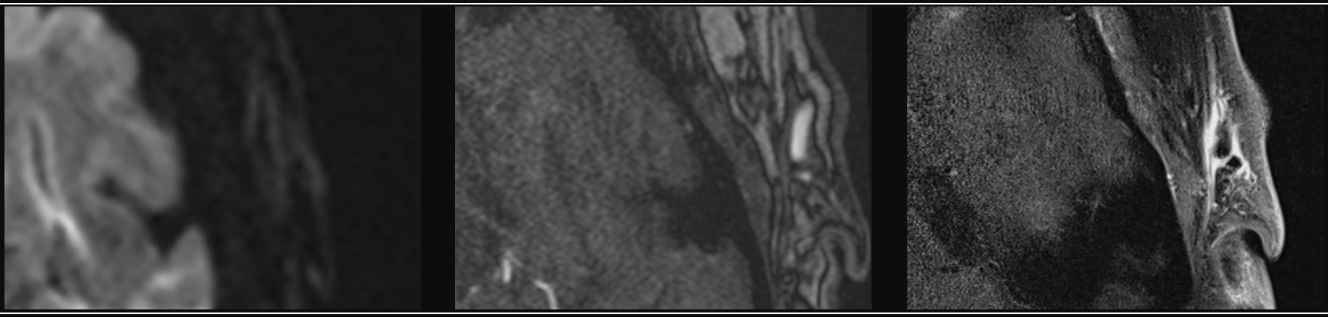

| DWI (left CSTA, score 2) | 3D-TOF-MRA (left CSTA) | T1-BB (left CSTA, score 3) |
|---|---|---|

#### Atlas Table 3. Examples of vertebral arteries with vasculitis in DWI

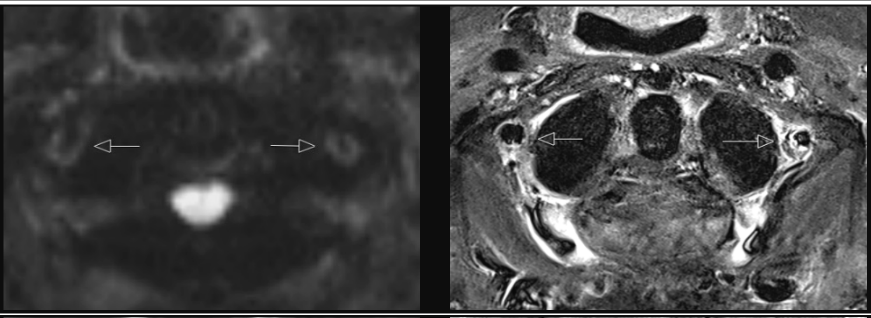

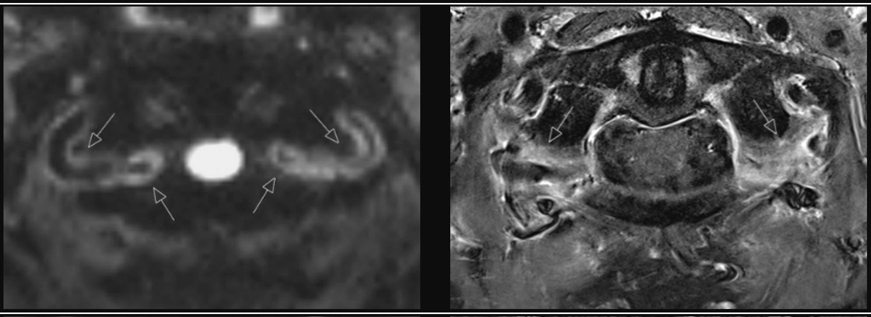

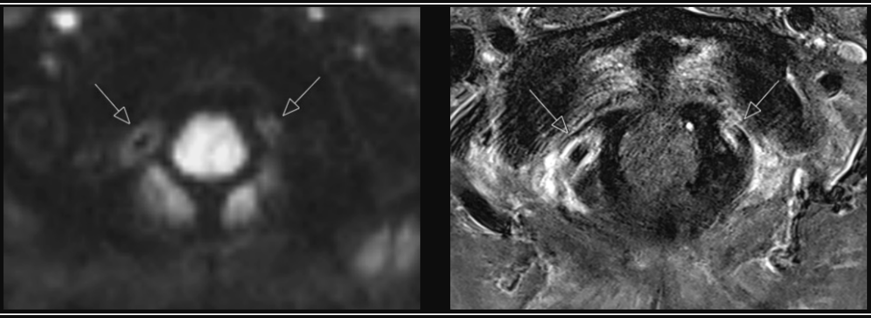

If the scan volume includes the V3 or V4 segments of the vertebral arteries, they should be screened for vasculitis. The V4 segment is usually visible only very faintly or not at all with DWI. The DWI- or T1-BB-scoring formally does not apply to arteries of this size. In this example, hyperintense signal can be seen on three consecutive slices in DWI with corresponding T1-BB slices with wall thickening and enhancement, more prominently on the left in the top two images. DWI may be used as a case-finding strategy for vasculitis of the vertebral arteries, especially if there are signs of stenosis on angiography.

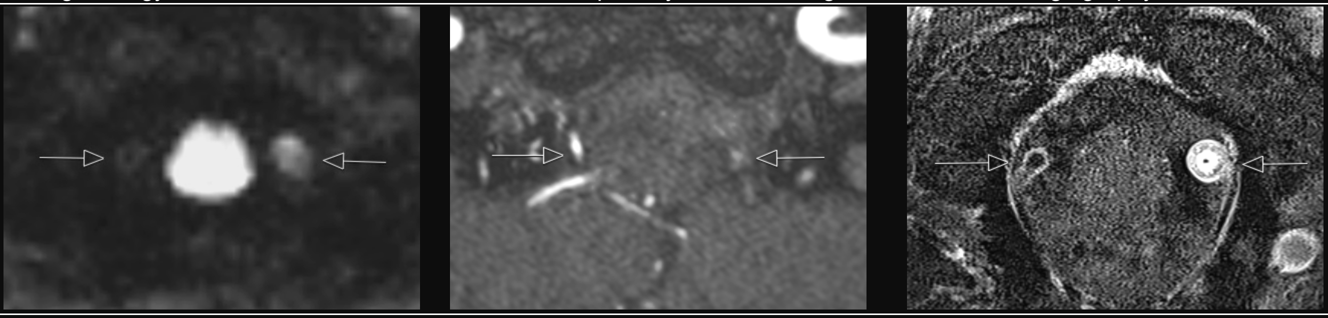

| DWI | 3D-TOF-MRA | T1-BB |
|---|---|---|

In this case of severe giant cell arteritis, bilateral vertebral arteries show abnormal hyperintense signal on DWI (right side borderline findings in DWI). In the arterial 3D-TOF-MRA the left vertebral artery shows only a very faint flow signal. On the corresponding T1-BB image, there is a very strong enhancement with severe vessel wall thickening of the left vertebral artery.

**Atlas Table 4. Pitfalls for DWI of the superficial cranial arteries**

**Lymph nodes**

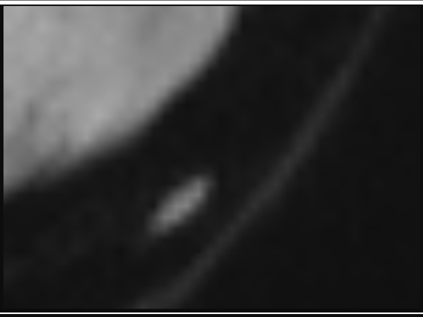

DWI (occipital left)

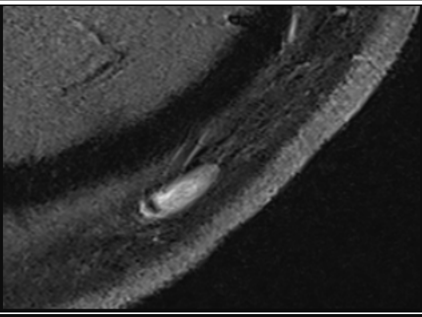

T1-BB (occipital left)

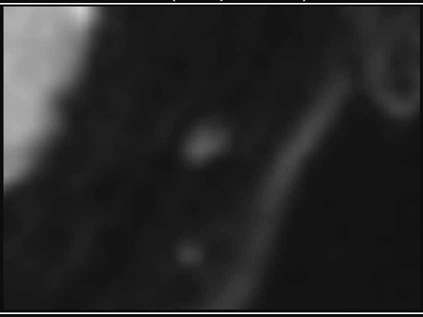

DWI (occipital left)

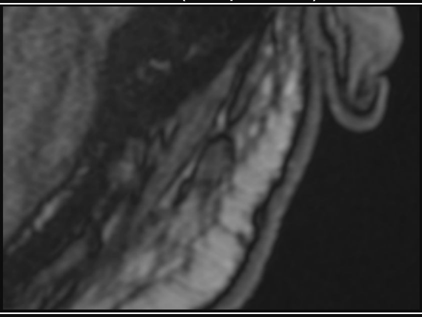

3D-TOF-MRA (occipital left)

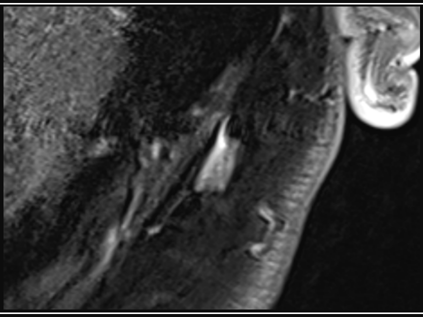

T1-BB (occipital left)

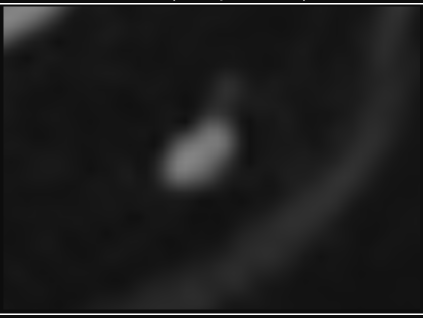

DWI (occipital left)

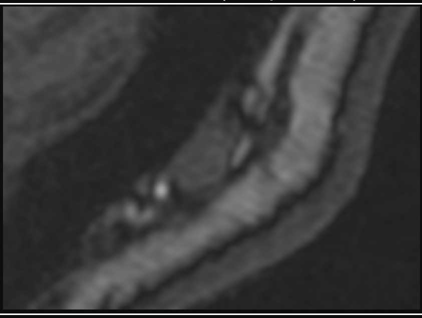

3D-TOF-MRA (occipital left)

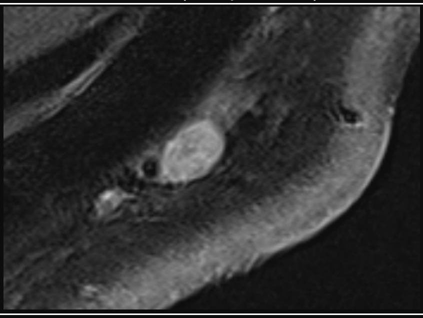

T1-BB (occipital left)

Lymph nodes typically show restricted diffusion with a bright signal on DWI images. Because a normal lymph node is not surrounded by edema, the demarcation from the surrounding fat is usually much sharper when compared to an artery with vasculitis. In addition, a normal lymph node is usually visible on only one or two, rarely three slices (4 - 5 mm slice thickness) and does not have the typical vascular course of an artery.

**Veins**

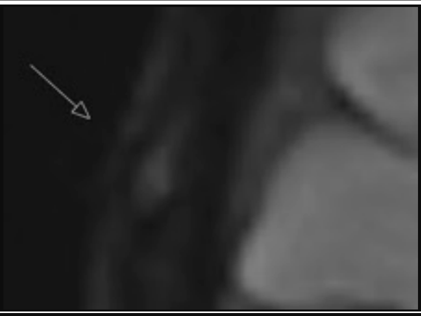

DWI (frontal right)

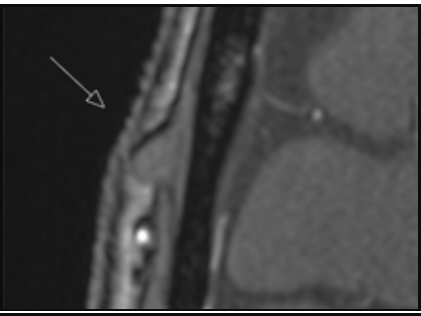

3D-TOF-MRA (frontal right)

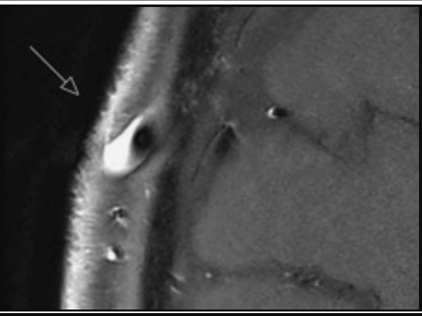

T1-BB (frontal right)

A vein (drop-shaped on T1-BB and 3D-TOF-MRA) has an abnormal DWI signal (formally score 2) with similar shape but no arterial signal in the 3D-TOF-MRA. Veins can rarely be detected on DWI images, especially if they have a large diameter.

| | | |
|---|---|---|
| 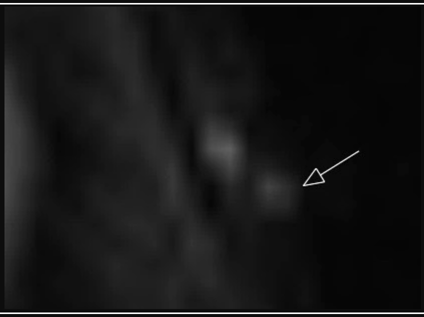 | 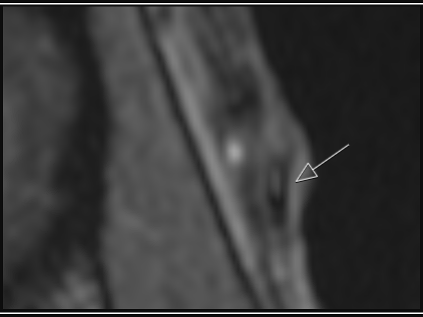 | 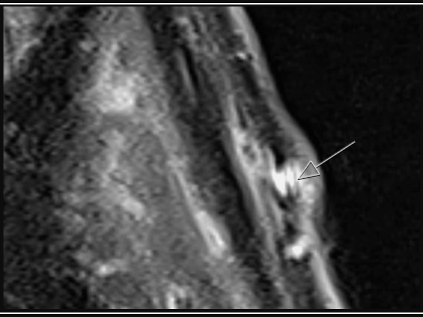 |
| DWI (frontal left) | 3D-TOF-MRA (frontal left) | T1-BB (frontal left) |
| A vein (arrow) crosses superficially to a frontal branch. The vein formally has a DWI score of 2, and the artery it crosses over has a DWI score of 3. In the 3D-TOF-MRA the vein shows no arterial signal. | | |
| **Parotid gland** | | |
| 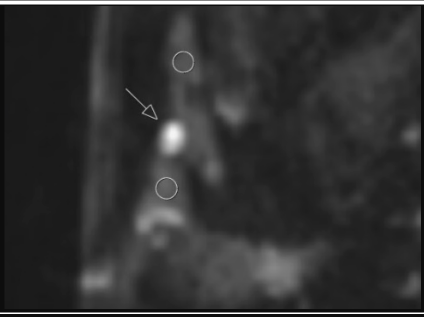 | 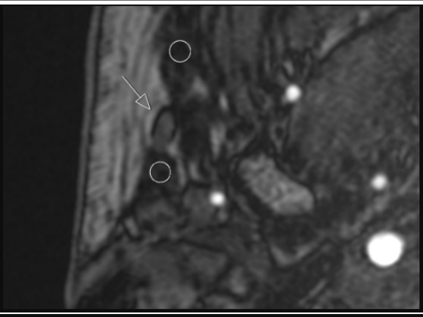 | 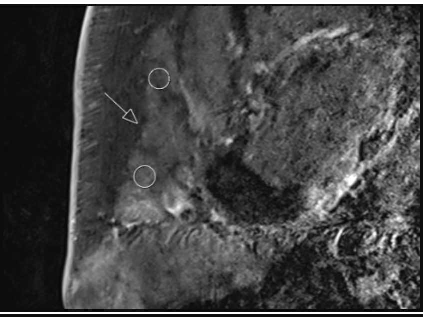 |
| DWI (right side) | 3D-TOF-MRA (right side) | T1-BB (right side) |
| On DWI images, the parotid glands (circles) can have variable signal intensities. The proximal temporal arteries can run right adjacent to the parotid glands. In DWI images it can be difficult to distinguish the signal of an artery with vasculitis from the signal of the parotid gland itself. The arrow shows a preauricular lymph node with its typical bright signal in DWI | | |
| **Muscle** | | |
| 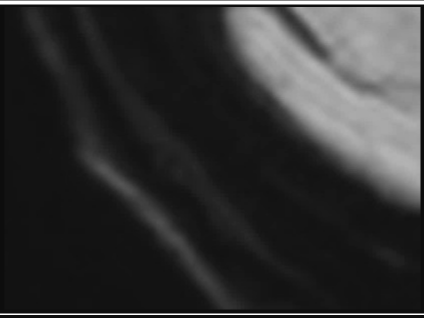 | 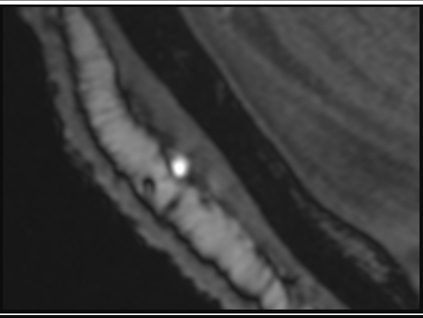 | 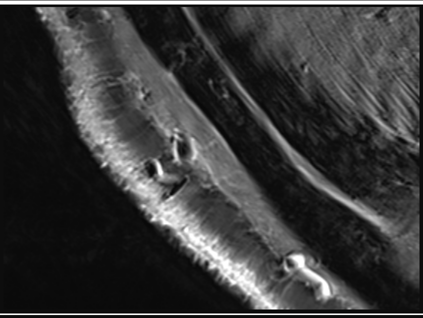 |
| DWI (occipital right) | 3D-TOF-MRA (occipital right) | T1-BB (occipital right) |
| 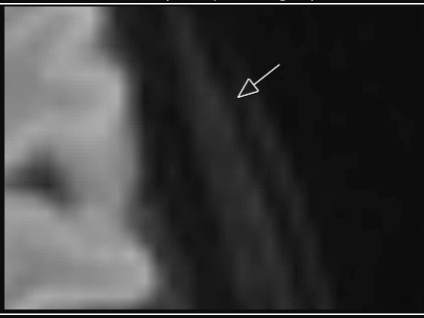 | 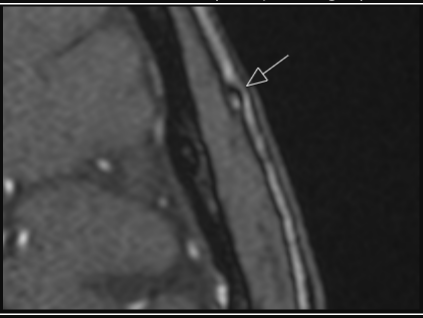 | 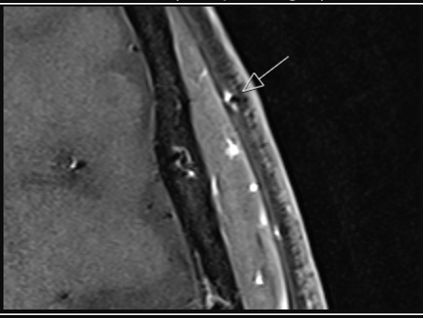 |
| DWI (frontal left) | 3D-TOF-MRA (frontal left) | T1-BB (frontal left) |
| On DWI images, muscle tissue usually shows a low intensity signal. Especially for the occipito-nuchal muscles (e.g., semispinalis capitis, occipital belly of the occipitofrontalis) or the periorbital muscles (e.g., orbicularis oculi), it is sometimes difficult to distinguish the muscular signal from the signal of arterial segments running in the axial plane or perivascular edema. The arrow indicates a left frontal branch. | | |

*Table 4 continued*

**Bone**

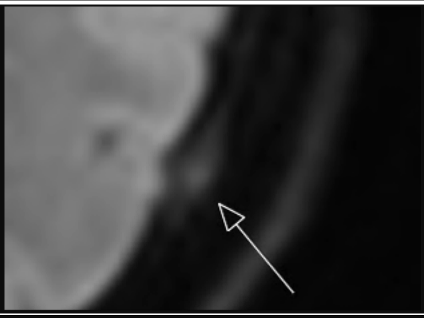

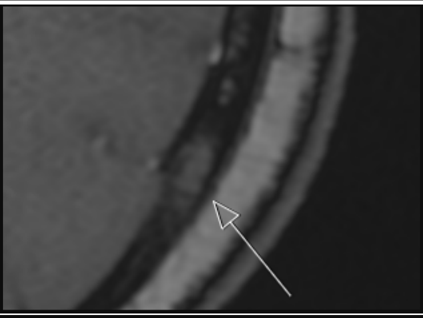

DWI (occipital left) | 3D-TOF-MRA (occipital left)

There is a prominent signal on the DWI image, but it projects onto the bone.

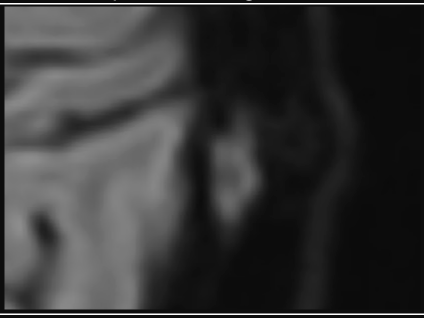

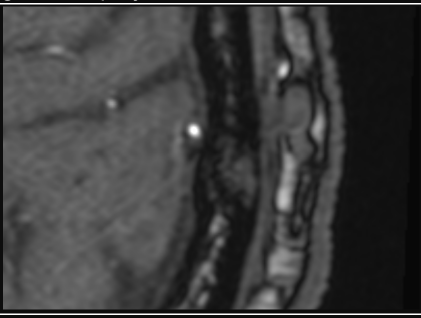

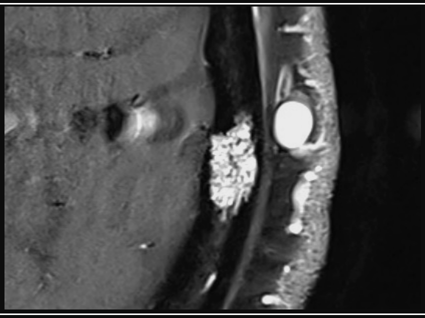

DWI (parietal left) | 3D-TOF-MRA (parietal left) | T1-BB (parietal left)

A bright signal projecting to the bone is seen on the DWI image with an intense corresponding T1-BB signal (hemangioma). If the structure is not in the subcutaneous fat, it should not be rated in DWI.

**Atlas Table 5.** **Pitfalls for T1-BB imaging of the superficial cranial arteries**

**Veins**

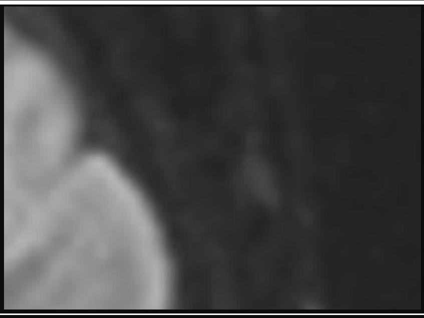

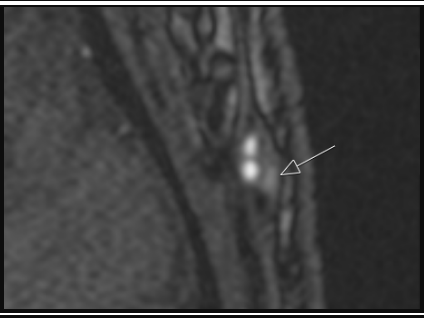

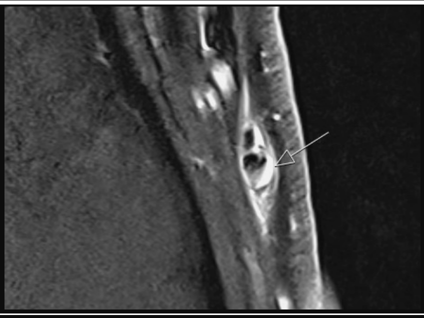

DWI (frontal left) | 3D-TOF-MRA (frontal left) | T1-BB (frontal left)

On the T1-BB image, a very bright signal corresponding to a small vein (arrows) can be seen superficial to the arterial lumen. This may lead to an incorrect estimate of the thickness and signal intensity of the arterial wall. In this case, vasculitis was nevertheless present, with a DWI score of 2 and a T1-BB score of 2-3.

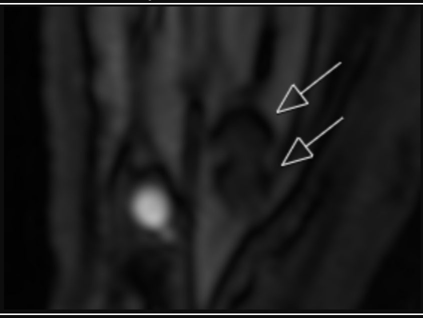

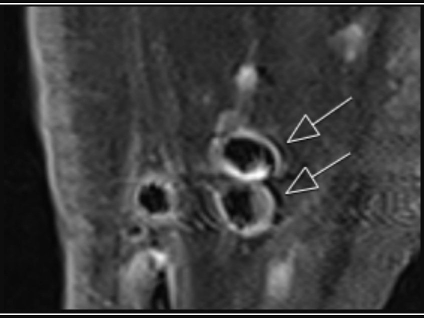

3D-TOF-MRA (CSTA right) | T1-BB (CSTA right)

It is relatively common for veins in the T1-BB to have a flow-void that simulates an artery. However, often the diameter of the vein is larger than the typical artery or the shape is not round. Comparison with the 3D-TOF-MRA can clearly identify the artery in such cases.

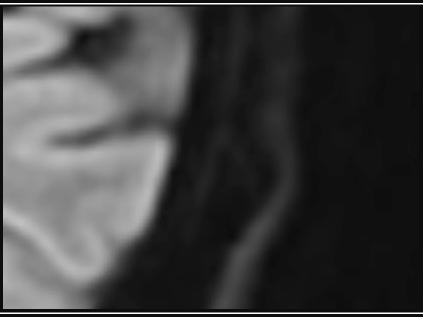

DWI (parietal left)

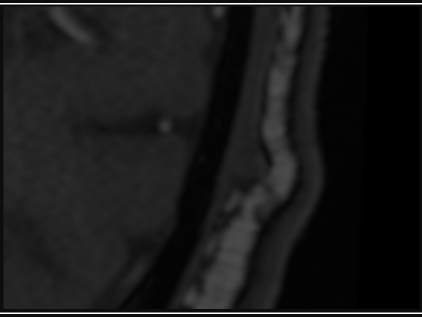

3D-TOF-MRA (parietal left)

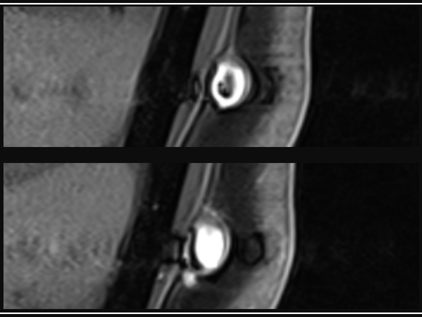

T1-BB (parietal left)

In this case, the vein has no flow signal on 3D-TOF-MRA and a DWI score of 0. On the T1-BB image, the vein appears to have a very thick wall simulating a vasculitis-like pattern. If this were true vessel wall enhancement, at least some degree of concomitant perivascular enhancement would be expected. A DWI score of 0 would also be very unusual. A few slices further caudally, no flow-void was present, which is the typical finding of veins in T1-BB.

**Occluded vessels**

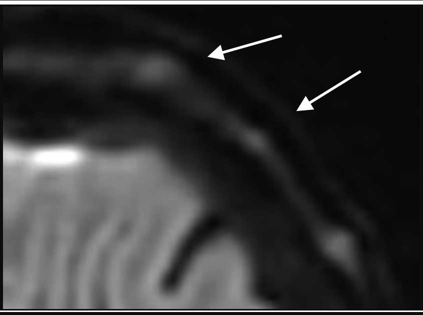

DWI (left frontal branch, score 3)

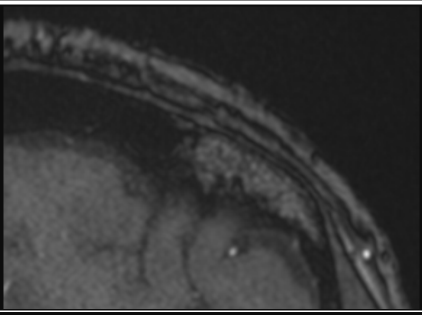

3D-TOF-MRA (left frontal branch)

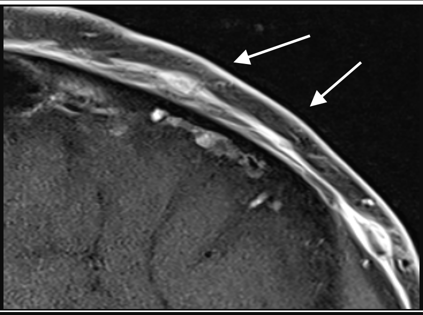

T1-BB (left frontal branch, score 3)

In the DWI image, three branches of the left frontal branch have a DWI score of 3. In the arterial 3D-TOF-MRA, only one of these branches showed an arterial flow signal; the two more medially located branches (arrows) did not, because they were occluded. Therefore, in T1-BB images, occluded arteries can easily be misinterpreted as veins because no flow-void is present. DWI can help identify occluded segments in severe vasculitis.

**Tangentially cut segments**

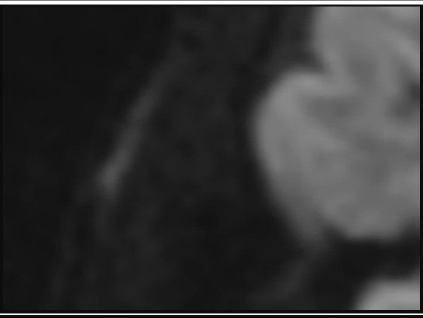

DWI (right frontal branch, score 3)

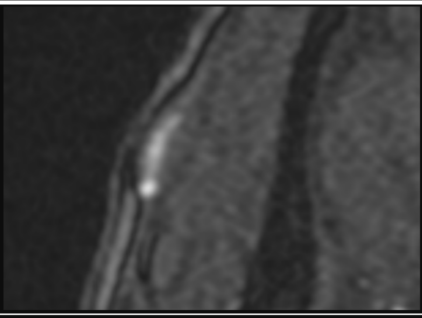

3D-TOF-MRA (right frontal branch)

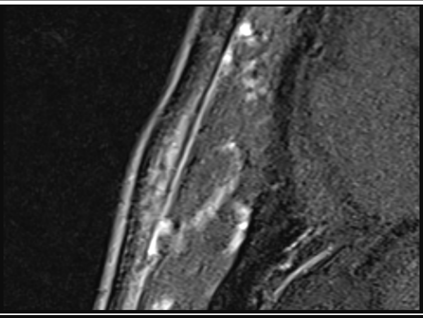

T1-BB (right frontal branch)

The course of the frontal branch can be almost parallel to the axial plane in certain parts. This regularly results in tangentially cut segments. In T1-BB, a lumen can be difficult to identify in such cases. Examination of the corresponding DWI image, can help in identifying a pathological T1-BB segment.

**Atlas Table 6. Change of DWI scores during follow-up and relapse**

**Case** **Timing**

**1**

**DWI**

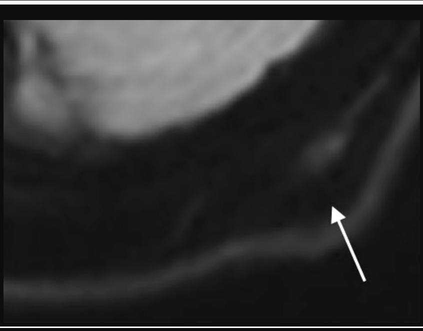

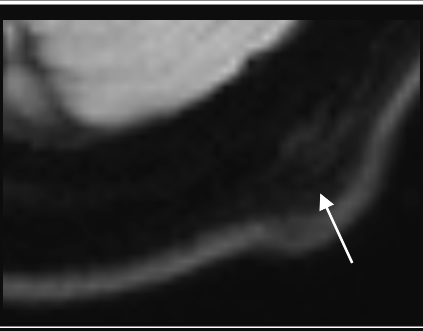

Baseline (DWI score 2) | Remission at 12 months (DWI score 1)

The left occipital artery has a score of 2 in both DWI and T1-BB images at baseline. After 12 months of treatment, the abnormal signal in DWI and T1-BB has resolved.

**T1-BB**

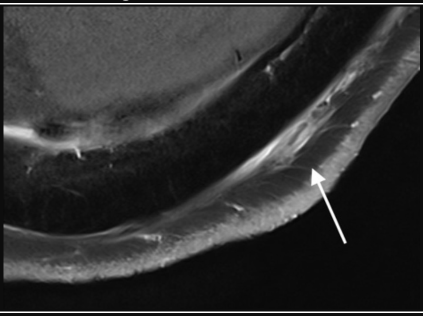

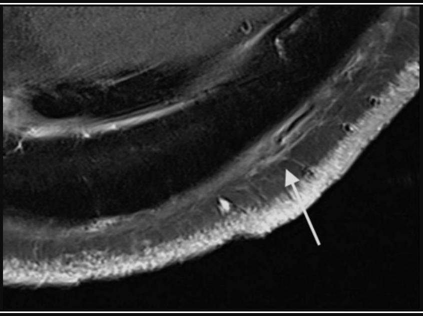

Baseline (T1-BB score 3) | Remission at 12 months (T1-BB score 1)

**2**

**DWI**

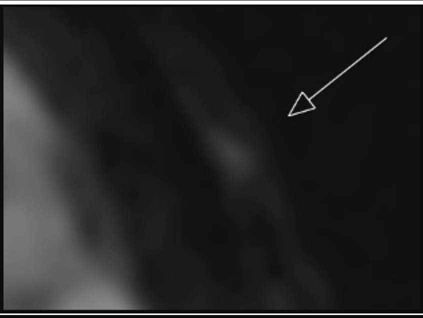

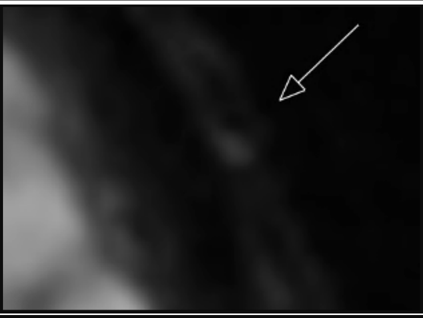

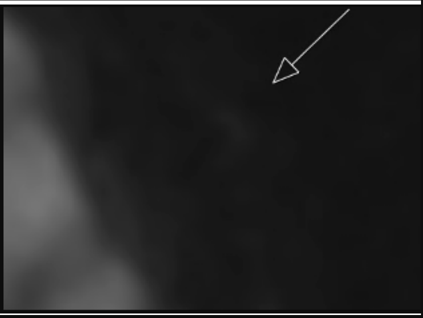

Baseline (DWI score 2) | Relapse at 6 months (DWI score 2) | Remission at 12 months (DWI score 0)

This patient relapsed after discontinuation of glucocorticoids and showed very similar findings of the frontal branch in DWI compared to baseline. After restarting glucocorticoids, a sustained clinical remission was achieved, and MRI showed normalization of the DWI signal.

**3**

**DWI**

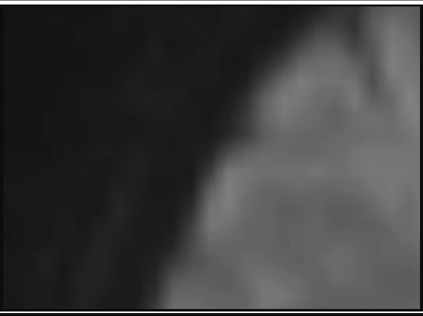

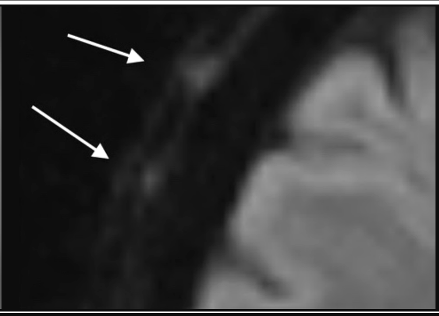

Baseline (DWI score 0) | Relapse at 24 months (DWI score 2 - 3)

MRI was normal at baseline in both the T1-BB and DWI sequences. Treatment was stopped after approximately 18 months of treatment in clinical remission. After a period of approximately 5 months without immunosuppressive therapy, the patient presented with a potential relapse. A repeat MRI scan now clearly showed affection of the superficial cranial arteries in both the DWI and T1-BB sequences.

**T1-BB**

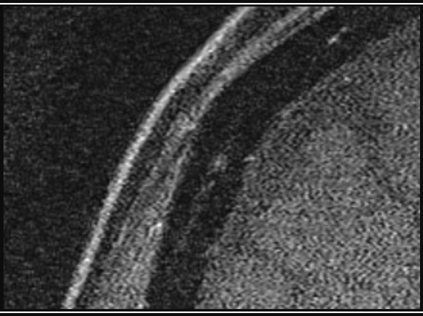

Baseline (T1-BB score 0)

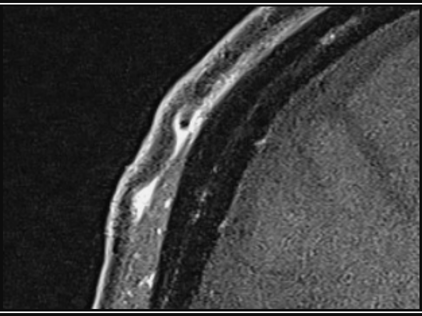

Relapse at 24 months (T1-BB score 3)

**4**

**DWI**

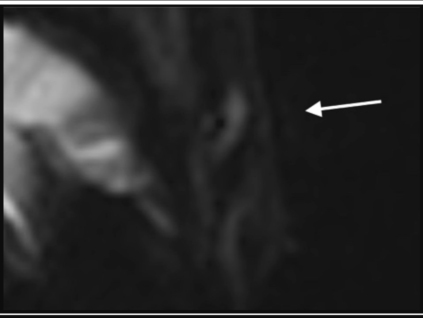

Baseline (DWI score 3)

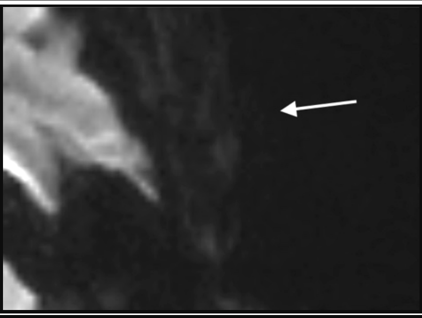

Remission at 12 months (DWI score 1)

The left CSTA is depicted as a linear structure in the DWI image at baseline. At 12 months, the artery is very difficult to delineate without direct comparison to a TOF-MRA or the T1-BB and the help of the crosshair.

**T1-BB**

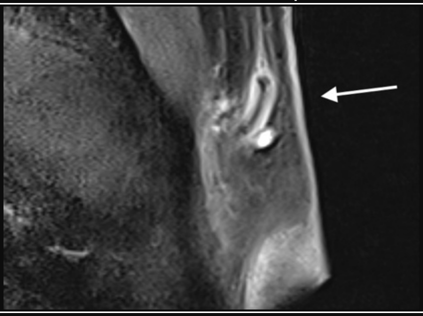

Baseline (T1-BB score 3)

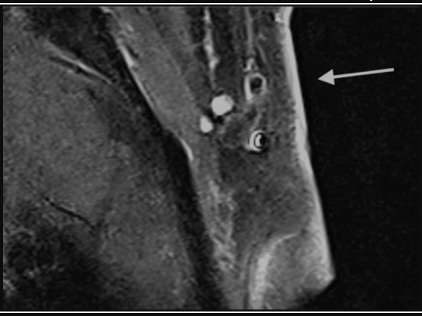

Remission at 12 months (T1-BB score 1)

**5**

**DWI**

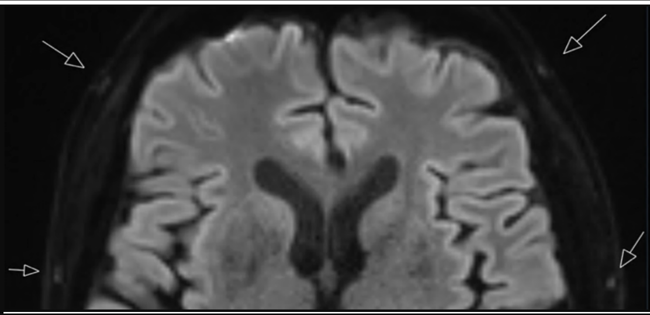

Baseline (DWI score 2 (right) – 3 (left))

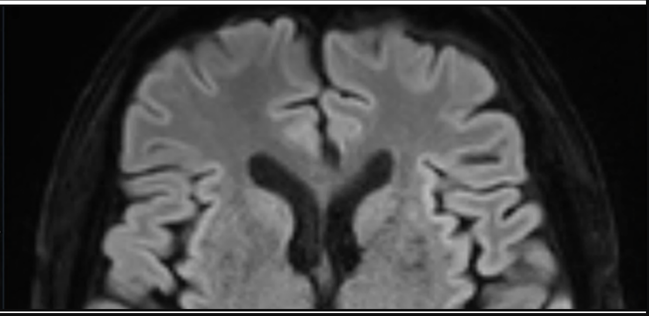

Remission at 12 months (DWI score 0 – 1)

The arteries with DWI scores 2-3 can easily be detected in the overview image of the frontal and parietal branches of the temporal arteries at baseline. At 12 months, no pathological signal can be detected (there were biopsies of the bilateral parietal branches between the two examinations)

**6**

**DWI**

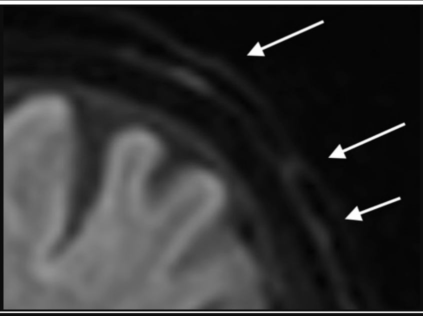

Baseline (DWI score 2 – 3)

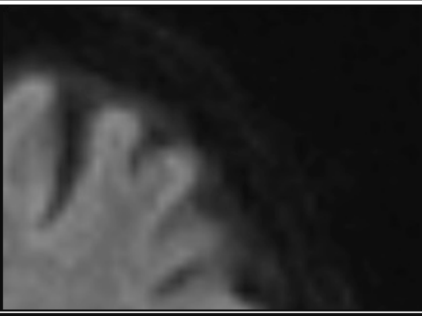

Remission at 12 months (DWI score 0)

At the level of the forehead, multiple branches with an abnormal DWI signal are seen adjacent to each other. Follow-up imaging shows complete regression with a score of 0 on both DWI and T1-BB images.

*Table 6 continued*

| | | |
|---|---|---|
| **T1-BB** | 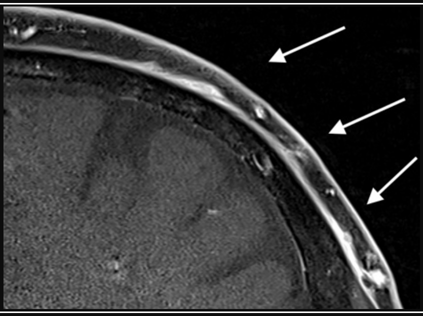 | 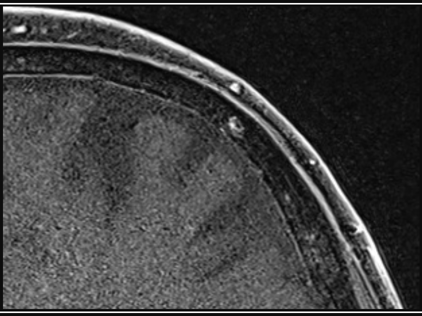 |
| | Baseline (T1-BB score 2 – 3) | Remission at 12 months (T1-BB score 0) |
| **7**<br>**DWI** | 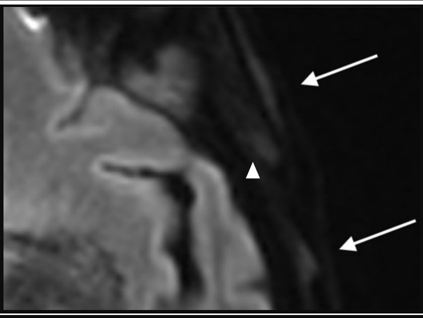 | 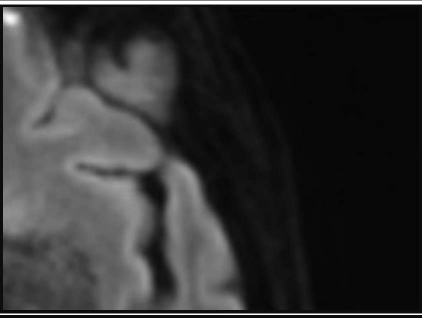 |
| | Baseline (DWI score 2 – 3) | Remission at 12 months (DWI score 0) |
| | In the T1-BB image at baseline, the lumen of the left frontal branch (upper arrow) is difficult to discern. The corresponding DWI image shows a linear hyperintense structure, typical of vasculitis affecting the frontal branch running superficial to the temporal muscle. In this example, there is a hyperintense area deep to the temporal fascia (arrowhead) within the muscle in DWI and T1-BB images. The finding is undetectable at 12 months, making it likely that it was also due to vasculitis. | |
| **T1-BB** | 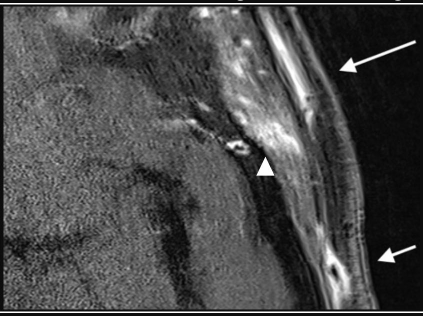 | 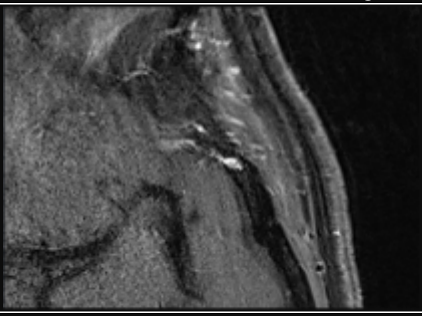 |
| | Baseline (T1-BB score 3) | Remission at 12 months (T1-BB score 0) |
